# Genome-Wide Study of Early Disability Severity in Multiple Sclerosis

**DOI:** 10.64898/2026.11.09.26367646

**Authors:** Soumeen Jin, Klementy Shchetynsky, Melissa Sorosina, Adil Harroud, Ali Manouchehrinia, Multiple MS, Lars Alfredsson, Jan Hillert, Maja Jagodic, Tomas Olsson, Fredrik Piehl, Federica Esposito, Ferdinando Clarelli, Mara Rocca, Massimo Filippi, Pernilla Stridh, Ingrid Kockum

## Abstract

**Background and Objectives:** Multiple sclerosis (MS) is a chronic neurological disorder characterized by demyelination and neuroaxonal injury. Genetics of MS susceptibility and long term disability is well characterized, but the genetic contribution to disability recorded earlier in the disease course is uncertain. We performed common-variant associations with the early period MS severity to identify potential causal loci, effector genes and associated pathways, and understand the relationships of early MS severity and other traits.

**Methods:** The primary outcome was the first age-related MS severity score (ARMSS), an age-adjusted severity measure, in 7,209 individuals of Swedish discovery cohorts and independent Italian and multi-national European replication cohorts. Replicated loci underwent conditional analysis, fine-mapping, time-to-event MS phenotype association analyses, and multi-omics-integrated gene prioritization. Using the genome-wide summary statistics, we also conducted heritability analyses, polygenic risk score association with MS susceptibility and late MS severity, gene-level analyses, cross-trait analyses, Mendelian randomization, and drug-repurposing screening.

**Results:** Two suggestive loci were replicated: rs1234317 on chromosome 1 (effect allele T: beta = 0.100, SE = 0.0189, P = 1.08 × 10^−7^) and rs929769 on chromosome 5 (effect allele T: beta = −0.0956, SE = 0.0184, P = 1.95 × 10^−7^). Fine-mapping supported rs929769 strongly in chromosome 5, and rs1234317 and rs6657136 in chromosome 1 with lower confidence. The SNPs did not show a significant change in effect sizes when adjusted for heritable traits and exhibited attenuated associations with long-term MS disability progression. *TNFSF4* was the top prioritized effector for rs1234317, *TNFSF18* for rs6657136, and *ADAMTS2* for rs929769. Early MS severity showed partial genetic overlap with MS susceptibility and late MS severity, but no significant correlation with a range of neurological, psychiatric, autoimmune, cognitive, and aging traits. Pathway analyses identified ion-channel regulation and folic acid metabolism. Mendelian randomization suggested nominal bidirectional causal associations with educational attainment. In silico drug screening nominated several candidate drugs.

**Discussion:** This study identifies candidate causal variants of early MS severity and their effector genes: *TNFSF4*, *TNFSF18*, and *ADAMTS2*. It provides insights into the genetic architecture and therapeutic measures for the early phase of MS severity.

## Introduction

Multiple sclerosis (MS) involves immune infiltration of the central nervous system (CNS), leading to neurological disability. Interplay between genetic and lifestyle factors shapes susceptibility to MS. The Human Leukocyte Antigen (HLA) region has the strongest genetic effect^1^ on disease susceptibility, whereas >200 additional non-HLA loci contribute modest to small effects.^2^ Although many genetic risk factors for MS susceptibility have been identified, disease severity remains largely underexplored.

Recently, the first genetic loci (rs10191329, P<5 × 10^−8^; rs149097173, P<5 × 10^−6^) were reported to be associated with MS severity, highlighting the role of CNS resilience in long-term disability progression. A higher cortical lesion rate and brain atrophy was also associated with rs10191329 in people with MS (pwMS).^3^ Despite this evidence, other studies have failed to replicate the rs10191329 association,^4,5^ which is likely due to the low statistical power.^6^ Notably, a study suggested that the effect of severity variants might differ over the disease period, reporting that rs10191329 had the greatest effect between 14 and 20 years after MS onset,^4^ implicating that severity loci other than rs10191329 and rs149097173 could have effects in the earlier period of MS. Since the existing genome-wide association study (GWAS) on MS severity focused on the late disease phase, we aimed to elucidate the genetic architecture of the disease severity of the early phase of MS. In addition to GWAS, we conducted fine-mapping for causal variants identification, association analyses with time-to-event MS phenotypes of the replicated SNPs (eFigure 1), integration of multi-omics gene prioritization, and tissue-specific gene expression and gene-set enrichment analyses. We further examined genetic correlations with various traits, assessed causal traits in relation to early MS severity, and potential therapeutic compounds. Given that treatment development supported by genetics significantly increases the potential of success,^7^ our findings offer mechanistic insights and help in patient care in the early period of MS disability progression.

**Figure 1.**
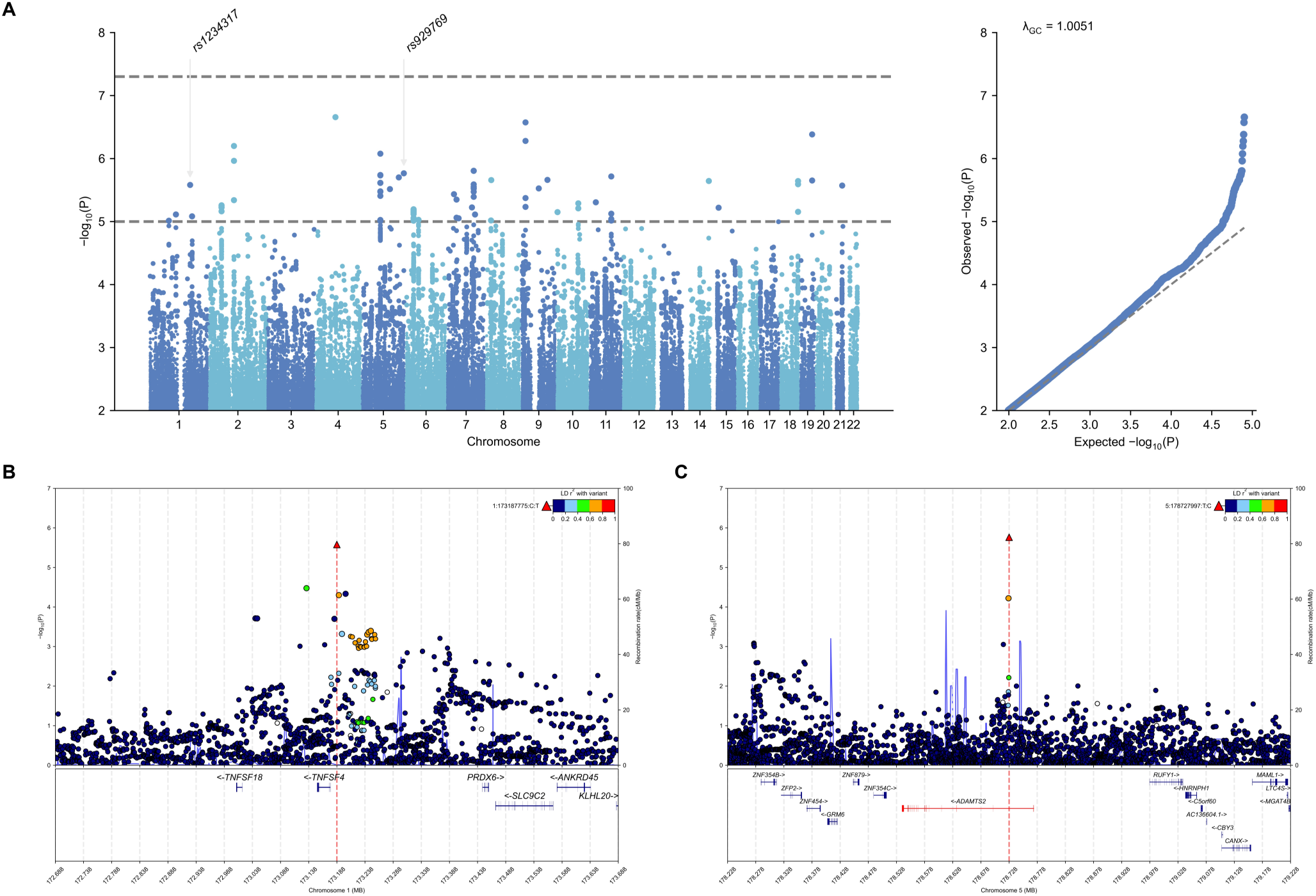
Genome-wide association study of early MS severity and the regional plots. (A) Manhattan and QQ plots for the discovery GWAS of early MS severity using the first ARMSS of pwMS. The horizontal dashed lines indicate the genome-wide significance threshold (P < 5 × 10^−8^) and the suggestive threshold (P < 1 × 10^−5^). The regional association plots for the lead SNPs, rs1234317 (B) and rs929769 (C), showing −log10(P) across ±500 kb around each lead variant. The SNP points are colored by linkage disequilibrium (r^2^) relationships based on 1000 Genomes European panel with the lead SNPs, and the recombination rates are shown in the background.

## Methods

### Standard Protocol Approvals, Registrations, and Patient Consents

The study setting, participant characteristics, eligibility to participate are presented in Table 1 and eSupplementary. The overview of this study is given in eFigure 1. Swedish patients have supplied written informed consent for the data collection and the study was approved by the local ethics committee. The MultipleMS prospective cohort study was approved by ethical review boards at nine participating sites. The Italian study was approved by the ethical committee of the IRCCS San Raffaele Hospital.

**Table 1.** Demographic characteristics of discovery and replication cohorts.

| **Variable** | **Discovery (N = 5,593)** |  |  | **Replication (N = 1,616)** |  | **Comparison** |  |
|---|---|---|---|---|---|---|---|
|  | **GSA<sup>d</sup>** | **OE<sup>d</sup>** | **GSA2<sup>d</sup>** | **OSR<sup>d</sup>** | **MMS<sup>d</sup>** | **P<sup>e</sup>** | **SMD** |
| **Sex** |  |  |  |  |  | **8.63 × 10<sup>−10</sup>** | **0.171** |
| Female | **1,265 (73.0%)** | **2,232 (73.9%)** | **563 (66.7%)** | **756 (65.2%)** | **289 (63.4%)** |  |  |
| Male | **467 (27.0%)** | **785 (26.0%)** | **281 (33.3%)** | **404 (34.8%)** | **167 (36.6%)** |  |  |
| **Disease course<sup>a</sup>** |  |  |  |  |  | **0.211** | **0.0404** |
| Bout onset | **1,715 (99.0%)** | **2,945 (97.6%)** | **840 (99.5%)** | **1,160 (100%)** | **398 (89.4%)** |  |  |
| Primary progressive | **17 (0.98%)** | **72 (2.39%)** | **4 (0.47%)** | **0 (0%)** | **19 (4.27%)** |  |  |
| Age at EDSS<sup>b</sup> | **35.6 (10.3)** | **41.5 (10.3)** | **36.3 (10.1)** | **34.2 (9.04)** | **34.1 (7.98)** | **3.33 × 10<sup>−69</sup>** | **0.48** |
| Age at onset | **31.1 (9.88)** | **34.5 (9.72)** | **32.1 (9.71)** | **30.0 (8.03)** | **32.3 (7.87)** | **6.65 × 10<sup>−23</sup>** | **0.268** |
| Disease duration<sup>b</sup> | **3.99 (4.38)** | **6.42 (5.15)** | **3.66 (4.39)** | **4.24 (4.47)** | **1.86 (2.87)** | **1.17 × 10<sup>−40</sup>** | **0.365** |
| Follow-up time<sup>c</sup> | **13.2 (6.90)** | **17.7 (7.32)** | **9.21 (7.32)** | **18.5 (8.00)** | **5.33 (3.20)** | **1.85 × 10<sup>−3</sup>** | **0.307** |
| EDSS<sup>b</sup> | **1.76 (1.56)** | **2.30 (1.89)** | **1.73 (1.45)** | **1.81 (1.07)** | **1.61 (1.18)** | **1.12 × 10<sup>−15</sup>** | **0.198** |
| ARMSS<sup>b</sup> | **4.59 (2.76)** | **4.91 (2.85)** | **4.59 (2.69)** | **4.13 (1.95)** | **3.73 (2.17)** | **1.82 × 10<sup>−32</sup>** | **0.307** |

a, the replication dataset had N = 1,577 for disease course. b, when repeated EDSS values were available, the reported age and score corresponded to the first recorded value. c, follow-up time was calculated from disease onset to the last EDSS measured. d, mean (SD) or n (%). e, chi-square test; Welch's t-test. P-values were two-sided and compared between the entire discovery and replication cohorts. For MMS, the time of the first symptom was used as a proxy for onset time. ARMSS, age-related multiple sclerosis severity score; EDSS, Expanded Disability Status Scale; SMD, standardized mean difference.

### Genome-wide association and meta-analyses

The genotype quality control (QC) and imputation resulting in three discovery Swedish chip cohorts (GSA, OE, GSA2) and two replication multi-national European chip cohorts (OSR, MMS) are provided in the eSupplementary and eFigure 2-4. To identify genetic variants associated with early severity of MS, we used a linear regression model implemented via PLINK 2.0.^8^ To ensure a normal distribution of the residuals, we applied a rank-based inverse normal transformation (RINT) on the first measured ARMSS. ARMSS is an age-adjusted MS severity score that ranks EDSS relative to pwMS in the same age bin (±2 years). EDSS is a neurological disability scale ranging from 0 to 10 to quantify MS disability.^9^ Year of birth, sex, and principal components (PCs) were included for adjustment, but disease-modifying therapy and smoking were not adjusted due to the possibility of collider biases.^10^ To evaluate potential residual confounding arising from population stratification or cryptic relatedness, we computed the genomic inflation factor and LDSC intercept using the HapMap3 variants and linkage disequilibrium (LD) scores from the 1000 Genomes Project phase 3.^11^ Lead variants were identified by iteratively extending *±*500*KB* around the most significant variant and merging the overlapping regions to the point where no variant below the *P <* 1 × 10^−5^ threshold were identified within the set window.^12^ Conditional analyses were performed to identify secondary signals. The lead variants were then tested for the replication analysis. Replication analysis was performed in the same regression model and the single-nucleotide polymorphisms (SNPs) were considered successfully replicated if the significance level of the one-sided P-value was below 0.05 with concordant direction of effect. The association statistics from the discovery and replication cohorts of the primary GWAS were meta-analyzed using a fixed-effects approach with METAL software.^13^

**Figure 2.**
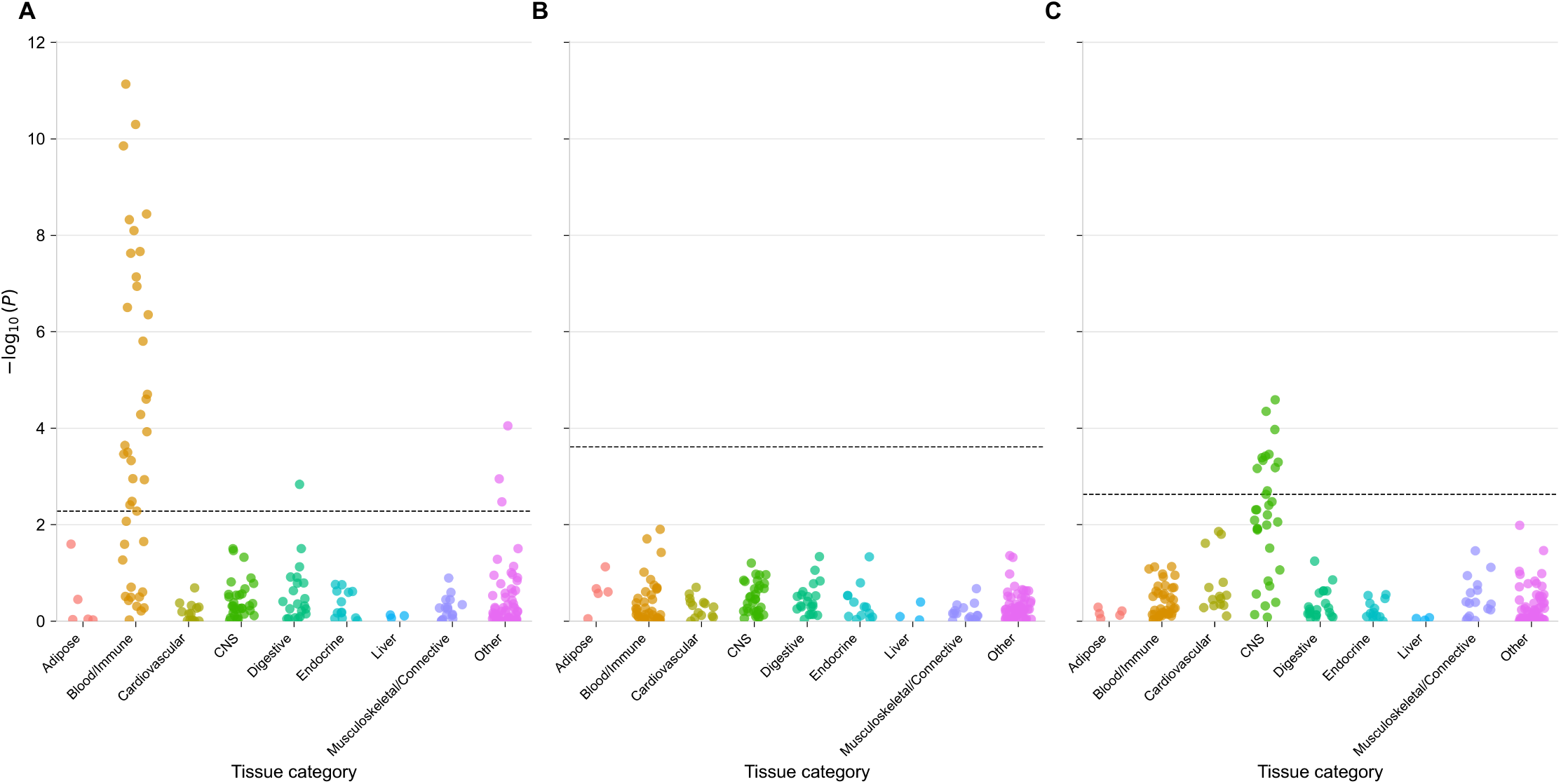
Heritability enrichment across tissues and cell types. The heritability enrichments were grouped into nine categories denoted on the X-axes. Y-axes convey -log10(P) of the enrichment coefficient. Heritability enrichments for (A) MS susceptibility,^2^ (B) early MS severity, and (C) late MS severity.^3^ The dashed lines indicate the Benjamini–Hochberg false discovery rate threshold ( < 0.05).

### Heritability estimation

To estimate SNP-based heritability, we used pooled genotype data of the discovery and replication populations and calculated a genomic relationship matrix (GRM) after cohort-level quality control and harmonization to a common autosomal biallelic SNP set.

Variants were trimmed with minor allele frequency (MAF) >= 0.01, genotype missingness <= 0.02 from the autosome. The related individuals were removed with the coefficients of relationship cutoff 0.025. We employed the resulting GRM to estimate SNP heritability using the restricted maximum likelihood method in GCTA (v 1.94.1).^14,15^ Given that SNP heritability is affected by LD and allele frequency,^16^ we also implemented a GREML-LDMS model incorporating ten GRMs constructed from variants categorized into five MAF bins (0.01–0.1, 0.1–0.2, 0.2–0.3, 0.3–0.4, and 0.4– 0.5), with each bin further subdivided based on the median LD score. Heritability analyses were adjusted for sex, year of birth, cohort, and ten PCs.

### Heritability enrichment analyses

We implemented stratified LDSC (version 1.0.1) to estimate SNP-based heritability enrichment in 53 baseline model functional annotations (version 1.2).^17^ To capture the tissue-specific contributions, we conducted stratified LDSC using a reference gene expression dataset of 205 tissues and cell types as given in LDSC.^17^ Multiple testing correction was carried out with false discovery rate (FDR)-adjusted *P <* 0.05. Prior to analysis, summary statistics were restricted to HapMap3 variants with imputation quality *R*^2^ *≥* 0.9 and excluded variants within the major histocompatibility complex (MHC) region.

### Fine mapping

We fine-mapped the two replicated loci using 1Mb windows around the lead variants. Summary statistics from the meta-analysis platforms were intersected with genotype data from all the cohorts. Platform-specific LD matrices were calculated after residualizing the genotypes for study-specific covariates and combined using sample-size weighting.

Kriging diagnostics and λs_rss were calculated to support the use of the weighted sum LD matrix. We utilized two methods for robustness: SuSiE-RSS^18^ and FINEMAP v1.4^19^ allowing up to 10 causal variants. We compared the SuSiE and FINEMAP results by estimating Pearson and Spearman correlation of posterior inclusion probabilities.

### rs1234317, rs6657136, and rs929769 associations with MS time-to-event phenotypes

The time-to-event disability analyses examined three traits: rate of Expanded Disability Status Scale (EDSS) progression over disease duration using a generalized linear mixed model (GLMM), time to clinically meaningful worsening (24-week confirmed disability worsening, Cox proportional hazards), and time to EDSS 6.0 with sensitivity analysis as time to sustained EDSS 6.0, both using interval-censored Cox model. Hazard ratios were calculated adjusted for sex, cohort, and PCs, with Bonferroni correction for multiple comparisons (*P* < 0.0083). Full model specifications are in eSupplementary.

### Sensitivity analyses

We first performed linear regression focusing on the SNPs of interest, without any transformation on the first ARMSS score. In addition, we utilized a linear mixed-model on RINT first ARMSS score using LDAK-KVIK,^20^ a genome-wide mixed-model tool with more realistic SNP effect size variation modeling than others.^21^ Both models were performed on discovery cohorts and meta-analyzed with METAL.^13^

We then respectively adjusted for age at onset, disease duration, and ever-smoking history in linear regression with RINT first ARMSS score, in a pooled discovery cohortand compared the estimates without the mentioned traits. Imputation chip and wave were accounted in each model. Collider bias was tested by examining the associations between each SNP of interest and the adjusted trait in linear regression framework.

### Multi-omics integrated gene prioritization and annotation

Candidate effector genes were prioritized using an integrative SNP-to-gene framework applied to the SNPs of interest and their LD proxies from the 1000 Genomes phase 3 European reference panel (r^2^ ≥ 0.6). Included strategies are 1) ANNOVAR/refGene positional variant-to-gene annotation, 2) molecular quantitative trait locus (QTL) evidence from expression QTL (eQTL)/splicing QTL (sQTL) resources, 3) chromatin interaction evidence from Hi-C, promoter-capture Hi-C (pcHi-C), and ABC resources, 4) Open Targets L2G predictions (L2G >= 0.5), 5) precomputed cS2G SNP-to-gene scores, 6) post-hoc transcriptome-wide association study (TWAS), and 7) colocalization evidence. Each strategy was weighted from the literature-prior evidence hierarchy. The gene that had the highest weighted sum of score for each SNP was prioritized as the candidate effector genes. For the annotations of the prioritized genes, tissue and cell-type expression profiles were examined using GTEx v10 bulk RNA-seq, the Human Protein Atlas single-cells and single-nucleus RNA-seq dataset. Full details of the resource references and the weighting scheme are in eSupplementary.

### Genome-wide gene association scan and pathway analyses

Two complementary genome-wide gene association scan (GWGAS) methods were employed: MAGMA v1.10 (linear, multi-model mode)^22^ and LDAK-KVIK-GBAT (mixed model).^20^ Pathway enrichment was assessed via MAGMA competitive gene-set analysis, Ingenuity canonical pathway and upstream regulator analyses, and FUMA tissue enrichment using GTEx v8 differentially expressed gene sets. Multiple testing correction used the Benjamini-Hochberg method. Full details are in eSupplementary.

### Genetic overlap of early MS severity with MS susceptibility, late severity, and non- MS traits

We tested 208 MS susceptibility variants^2^ for association with early MS severity using Bonferroni correction (*P* = 2.40 × 10⁻⁴). LD between early MS severity (rs1234317, rs6657136, and rs929769) and susceptibility SNPs were calculated using PLINK 2.0.^8^ The concordance in effect directions was evaluated with an exact binomial test.

Polygenic scores (PGS) were constructed with respect to the MS susceptibility variants and early MS severity were regressed on the PGS in GSA2 chip cohort two ways, one with PGS, year of birth, sex, and the PCs, and another without PGS and only the covariates. GSA2 chip cohort was utilized since the participants do not overlap with MS susceptibility GWAS.^2^ Incremental RZ was calculated from the R^2^ difference of the two.

For the genetic comparison between early and long-term MS severity, we tested rs10191329 and rs149097173 from the late MS severity GWAS^3^ in association with early MS severity. LD with early severity SNPs was calculated using PLINK 2.0.^8^ PGS with respect to suggestively significant late severity SNPs (*P* < 5 × 10⁻⁶) were constructed after LD clumping (r^2^ < 0.01) and incremental RZ was calculated in the same manner in the GSA2 chip cohort. GSA2 chip cohort was selected due to non-overlap of the participants.

For genetic comparison with non-MS traits, we evaluated phenome-wide associations for our SNPs of interest across the GWAS Catalog, UK Biobank, and FinnGen. Genome-wide cross-trait analyses were performed using LDSC for 16 neurological, psychiatric, autoimmune, cognitive, and aging traits. Full details and references for the implementations are in the eSupplementary.

### Mendelian Randomization

Two-sample MR was performed with four exposures: 25-hydroxyvitamin D, body mass index (BMI), lifetime smoking index, and educational attainment. Instruments were selected at two P-value thresholds (*P* < 5 × 10⁻⁸ and *P* < 1 × 10⁻⁵) after LD clumping (rZ < 0.001). Primary analysis used inverse-variance weighted (IVW) with random effects; sensitivity analyses included MR-RAPS, weighted median, MR-PRESSO, and MR-Egger. Reverse MR was also performed. Full details are in the eSupplementary.

### Genomics-based drug discovery

In silico drug prioritization leveraged two complementary strategies: 1) compound-gene matching via the Scalable Precision Medicine Open Knowledge Engine (SPOKE) graph database, querying ChEMBL, BindingDB, PharmGKB, DrugBank, and Therapeutic Target Database for compounds targeting the top candidate effector genes and the LDAK-KVIK-GBAT FDR-significant genes; and 2) TransPhar analysis to identify compounds with inverse transcriptional correlation between the predicted genetically regulated expression profiles of early MS severity and the compound-regulated gene expression in LINCS Cmap L1000 data. Full details are in the eSupplementary.

## Results

### Heritability analyses of early MS severity

We analyzed 7,209 pwMS European descent, with the discovery cohort *(N* = 5,593) having a mean age of 38.9 years at first EDSS assessment and a mean disease duration of 5.25 years, while the replication cohort *(N* = 1,616) had corresponding values of 34.2 and 3.57 years (Table 1). For the single-component heritability analysis, the estimates were imprecise (h^2^= 0.0388, SE = 0.0658), yet in multi-component model, the analysis gave estimates in better precision (h^2^ = 0.185, SE = 0.0947, 95% CI = [-0.0006, 0.3706]; eTable 1). The heritability partitioning across the 53 functional genomic annotation categories (eTable 2) nor for the gene sets across 205 tissues and cell types (eTable 3) did not find any enrichment.

### Genome-wide association study and fine-mapping

To understand the genetic components of the disability severity in the early period of MS, we conducted GWAS using rank-inverse normalized first ARMSS scores,^23^ in a linear regression framework in each chip cohort, followed by meta-analyses. The genomic inflation was modest (_GC_ = 1.01) and the linkage disequilibrium score regression (LDSC) intercept was not significantly different from 1 (estimate = 1.00, *SE* = 0.00632). We identified 33 independent SNPs with suggestive significance (*P* < 1.0 × 10^−5^, Fig. 1A), and of these SNPs, rs1234317 (1:173187775:C:T) and rs929769 (5:178727997:T:C), were replicated (Table 2, eTable 4). Conditional analyses did not reveal additional signals in each locus (Fig. 1B and 1C).

**Table 2.** Summary statistics of replicated causal candidate SNPs on early MS severity.

|  |  |  | **Discovery** |  |  |  |  | **Replication** |  |  |  |  | **Meta-analysis** |  |  |  |  |
|---|---|---|---|---|---|---|---|---|---|---|---|---|---|---|---|---|---|
| **Marker** | **A1** | **A2** | **Freq1** | **β (SE)** | **P** | **Het P** | **N** | **Freq1** | **β (SE)** | **P** | **Het P** | **N** | **Freq1** | **β (SE)** | **P** | **Het P** | **N** |
| 1:173187775:C:T (rs1234317) | **T** | **C** | **0.2530** | **0.1001 (0.0213)** | **2.63 × 10<sup>−6</sup>** | **0.5515** | **5,591** | **0.2315** | **0.1014 (0.0409)** | **0.01308** | **0.9879** | **1,607** | **0.2484** | **0.1004 (0.0189)** | **1.079 × 10<sup>−7</sup>** | **0.8796** | **7,198** |
| 1:173133986:A:G (rs6657136) | **A** | **G** | **0.3803** | **0.0812 (0.0196)** | **3.349 × 10<sup>−5</sup>** | **0.708** | **5,591** | **0.3230** | **0.1338 (0.0385)** | **0.0005126** | **0.5669** | **1,554** | **0.3685** | **0.0920 (0.0175)** | **1.346 × 10<sup>−7</sup>** | **0.6451** | **7,145** |
| 5:178727997:T:C (rs929769) | **T** | **C** | **0.5273** | **−0.0973 (0.0203)** | **1.72 × 10<sup>−6</sup>** | **0.2253** | **5,591** | **0.5464** | **−0.0879 (0.0427)** | **0.03949** | **0.2009** | **1,338** | **0.5309** | **−0.0956 (0.0184)** | **1.947 × 10<sup>−7</sup>** | **0.3245** | **6,929** |

A1, effect allele; A2, non-effect allele; Freq1, frequency of A1. β (SE) denotes the per-allele effect estimate and standard error. P-values test β = 0; Het P is the within-analysis heterogeneity test. Discovery used Swedish cohorts, replication used Italian and MultipleMS cohorts.

Fine-mapping with SuSiE-RSS and FINEMAP prioritized rs929769 at chromosome 5 as a single-variant 95% credible set (SuSiE-RSS posterior inclusion probability (PIP) = 0.964, FINEMAP posterior probability (PP) = 0.643). At chromosome 1, the primary 95% SuSiE credible set had two SNPs with the probabilities of 0.561 (rs6657136) and 0.399 (rs1234317), while the secondary set contained 1,399 variants including rs6657136 and rs1234317. FINEMAP assigned two-causal-variant model as the most probable in this locus (PP = 0.620), yet the highest individual two-variant configuration had PP=0.009. Detailed LD-matrix diagnostics, model comparisons, and SNP-level posterior probabilities are provided in eFigure 5 and eTables 5-6.

In the meta-analyses of the three SNPs across the discovery and replication cohorts, rs929769_T allele decreased the early MS severity (β= -0.0956, SE= 0.0184, P=1.95 × 10^−7^), rs1234317_T allele increased it (β= 0.100, SE= 0.0189, P=1.08 × 10^−7^), and rs6657136_A allele increased the severity (β= 0.0812, SE= 0.0175, P=1.35 × 10^−7^; Table 2).

### Sensitivity analyses of rs929769, rs1234317, and rs6657136

We then tested each SNP in linear mixed models and did not find notable difference in effect size or statistical significance compared to main analysis. We also conducted linear regression without RINT for clinical interpretability, which exhibited significance in effects (eTable 7). In addition, for these three SNPs we adjusted for several traits: age at onset, disease duration at first EDSS, and ever-smoking history to detect any confounding effects. No evidence of collider biases was observed and all three SNPs remained significant and similar in effect sizes after adjustments (eTable 8, eFigure 6).

### rs1234317, rs6657136, and rs929769 associations with MS time-to-event phenotypes

We next examined whether the prioritized SNPs affect MS progression using three disability accumulation MS phenotypes: rate of EDSS progression, time to 24-week confirmed worsening, and time to EDSS 6 with interval censoring. Time to sustained EDSS 6 was a sensitivity analysis of time to EDSS 6. Overall, associations with MS disability accumulations were weaker and less consistent. The rs1234317_T allele was associated with shorter time to EDSS ≥ 6 (HR = 1.15, SE = 0.0504, P = 0.00688), and rs929769_C was associated with higher EDSS scores over disease duration (β = 0.159, SE = 0.0472, P = 0.0007). No variant showed consistent evidence of altered EDSS progression rate or 24-week confirmed disability worsening. Full per-variant analyses of the tested MS progression phenotypes are provided in eFigures 7-9 and eSuppplementary.

### Multi-omics integrated gene prioritization

Integrative SNP-to-gene prioritization of 23 LD proxies for the three candidate causal variants are shown in eTable 9 and were used for prioritizing the potential effector genes. TNFSF4 was prioritized for rs1234317 based on positional, cS2G, and L2G evidence. The bulk and single-cell RNA-seq annotations for TNFSF4 supported its expression in immune and CNS-relevant cell types. TNFSF18 was prioritized for rs6657136 in positional, eQTL, and chromatin-interaction mapping. The TNFSF18 expression was found across brain tissues and neuronal, macrophage/microglial, and glial populations. ADAMTS2 was prioritized for rs929769 based on positional and cS2G evidence. Its expression was seen in fibroblasts, macrophages, oligodendroglial cells, and neuronal clusters. Detailed gene-prioritization scores, expression profiles, eQTL results, and chromatin-interaction annotations are in eSupplementary text, eTable 9, and eFigures 10-12.

### Genome-wide gene association scan and pathway analyses

Genome-wide gene-association scan and pathway analyses provided suggestive support for biological pathways relevant to early MS severity. Although MAGMA did not identify FDR-significant genes, LDAK-KVIK-GBAT identified 189 FDR-significant genes associated with early MS severity (eFigure 13, eTables 10-11). Tissue-specific expression analyses demonstrated enrichment of the FDR-significant genes among downregulated differentially expressed genes (DEGs) in brain regions, upregulated DEGs in EBV-transformed lymphocytes, and bidirectional DEGs in the left ventricle (eFigures 14-15, eTables 12-13). Pathway analyses highlighted sinoatrial node cell development and differentiation, cardiac pacemaker cell differentiation, folic acid transport, and folic acid binding (eTables 14-18, eFigure 16). The full results are in eSupplementary.

### Genetic relationships of early MS severity with MS susceptibility, late MS severity, and non-MS traits

Genetic overlap analyses suggested partial but limited sharing between early MS severity, MS susceptibility, and late MS severity. Compared with published MS susceptibility ^2^ and late-severity GWAS results ^3^, early MS severity showed no significant immune- or CNS-cell heritability enrichment pattern matching those traits (Figure 2; eTable 3). One MS susceptibility variant, rs4409785 (eTable 19), was associated with early MS severity after Bonferroni correction (*P*=0.0302). The proportion of susceptibility variants in concordant direction of effect with the early severity variants significantly differed from randomness (*P* = 0.0266, eTable 20). The late-severity variant rs10191329 was associated with early MS severity in the same direction of effect (β =0.0623, *P* = 0.00588), although in cohorts that did not overlap with the late MS severity GWAS,^3^ the association was insignificant (β =0.0327, *P*=0.546). The LDs between the early MS severity loci and the published susceptibility or late-severity variants were low (susceptibility: *r*^2^ ≤ 0.00133; late-severity: *r*^2^ ≤ 2.99 × 10^−4^; eTables 21, 22; eFigure s17A, 17B), and PGS for MS susceptibility and late severity explained small variance in early MS severity (susceptibility: *ΔR*^2^ = 0.00343; late-severity *ΔR*^2^ = 0.00211; eTables 23, 24). Phenome-wide lookup of rs929769, rs6657136, and rs1234317 found no significant evidence of pleiotropic associations and genome-wide cross-trait analyses reported limited shared genetic contributions of early MS severity with broader neurological, psychiatric, cognitive, autoimmune, or aging-related traits (eTable 26; eFigure 18). The detailed results are in eSupplementary.

### Mendelian randomization of modifiable traits

In two-sample MR analyses, we found no evidence of causal or reverse-causal associations for 25-hydroxyvitamin D, body mass index, or lifetime smoking index, although the reverse analysis for lifetime smoking showed heterogeneity (P = 5.3 × 10^-4^). Higher educational attainment suggested a nominal inverse causal association with early MS severity across the MR estimators, with no evidence of directional pleiotropy by Egger intercept (P = 0.732 and 0.346) or MR-PRESSO global tests (P = 0.201 and 0.091). Reverse MR for educational attainment also suggested a nominal causal association (β = -0.0755, SE = 0.0327, P = 0.0209), with acceptable heterogeneity (P = 0.253; Figure 3; eTables 27-28).

**Figure 3.**
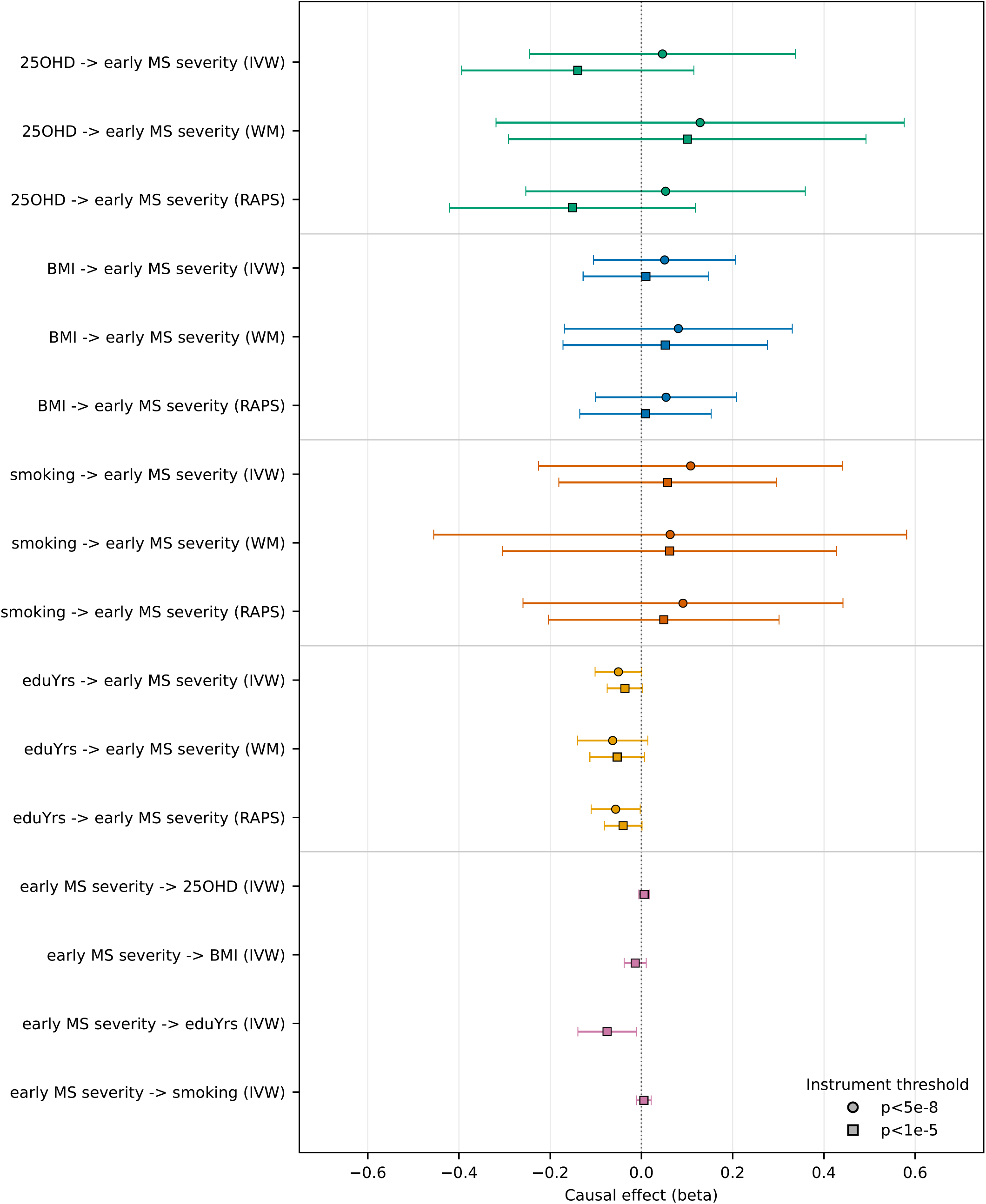
Mendelian randomization of four traits and early MS severity. Forest plots demonstrate the Mendelian randomization (MR) causal estimates with 95% confidence intervals across multiple MR methods between 25-hydroxyvitamin D level, BMI, years of education, and lifetime smoking index, and early MS severity. The arrows indicate the causal direction. 25OHD, 25-hydroxyvitamin D level; BMI, body mass index; eduYrs, years of education; smoking, lifetime smoking index; IVW, inverse-variance weighted; WM, weighted median; RAPS, robust adjusted profile score

### In silico drug screening

In silico drug-prioritization analyses, compound-gene matching nominated candidate compounds targeting TNFSF4, TNFSF18, ADAMTS2, and genes identified in gene-based analyses. However, Inverse correlation analysis between the predicted genetically regulated expression (GREx) and LINCS L1000 gene expression signature did not identify any compounds (eTables 29-31; eFigure 19). The detailed description of the results are in eSupplementary.

## Discussion

We conducted a GWAS of early MS severity using the first ARMSS measurement to characterize disability at an early stage when long-term progression and cumulative treatment exposure have not dominated the phenotype. Disability accumulation in MS involves two partially separable processes: relapse-associated worsening (RAW) that predominates early, and progression independent of relapse activity (PIRA) that contributes increasingly as the disease evolves.^24^ Our study design leveraged early ARMSS and excluded measurements proximal to relapses, to reduce the relapse-driven disability dominance but retain sensitivity to early PIRA-related disability accrual. We identified two replicated loci, at chromosomes 1 and 5, and fine-mapping prioritized rs1234317, rs6657136, and rs929769 as candidate causal variants. Integrative gene prioritization nominated *TNFSF4*, *TNFSF18*, and *ADAMTS2* . We also observed partial overlap of early MS severity genetics with MS susceptibility and less with late severity, but limited overlap with broader disease-related traits. Potential bidirectional causal relationship was indicated between early MS severity and the education years, and several pathways and drug candidates were suggested.

Although SNP-heritability estimates was barely crossing the borderline null value in 95% CI, the results imply common-variant contribution to early MS severity, which could be detected with larger participant size. Fine-mapping provided the strongest support for rs929769 at chromosome 5, whereas chromosome 1 locus demonstrated rs6657136 and rs1234317 as the most probable causal SNPs yet with lower confidence shown by large number of variants in the second credible set. Sensitivity analyses confirmed the effect of all three SNPs. The attenuated associations of these variants with later time-to-event disability outcomes suggest that their effects may be more relevant to early severity than to long-term disability accumulation.

Integrative multi-omics-based candidate effector gene investigation prioritized *TNFSF4*, *TNFSF18*, and *ADAMTS2*. *TNFSF4* encodes OX40L, a regulator of T-cell activation. Astrocytic OX40L can stimulate OX40-positive T cells in CNS tissue, promoting glial activation, blood–brain barrier (BBB) breakdown, and axonal damage.^25–27^ *TNFSF18* encodes glucocorticoid-induced TNFR-related protein (GITR) ligand (GITRL) expressed in immune and CNS-resident cell types and participates in regulatory T-cell expansion, CNS neuroinflammatory signaling.^28,29^ *ADAMTS2* encodes a secreted metalloprotease involved in extracellular matrix (ECM) remodeling, with expression localized to excitatory neurons and perivascular fibroblast-like cells. The dysregulation in ADAMTS family members are associated with ECM degradation and MS progression,^30–32^ suggesting the possible role of ADAMTS2 in similar mechanistic context.

Genetic comparison with MS susceptibility and late MS severity suggested partial but limited overlap. Although the selected susceptibility and late-severity variants were associated with early MS severity and non-random concordance in effect direction between early severity and MS susceptibility was detected, early severity did not show the immune-cell or CNS-cell heritability enrichment patterns observed for MS susceptibility and late severity, respectively. This could be due to early severity not being driven by either mechanism or power issue.^2,3^ However, early severity regressed on PGS constructed by the two phenotypes explained little additional variance. Collectively, these findings suggest that early MS severity may share selected genetic components with MS susceptibility and later disability severity, but to limited extent.

In examining the genetic relationships of early MS severity with various phenotypes, neither phenome-wide analyses for rs929769, rs6657136, and rs1234317, nor genome-wide genetic correlations with neurological, psychiatric, aging, and autoimmune traits revealed significant associations. The results could be due to early severity having unique genetic architecture or limitation in heritability estimates due to available sample size.^33^

Secondary overrepresentation analyses of early MS severity GWGAS genes in tissues and cell-specific DEGs may reflect shared transcriptional characteristics with reduced expression in brain tissues, active expression in immune cells, and cellular energy metabolism or stress response signatures across excitable cell types. Pathway analyses highlighted non-canonical MS pathways that may reflect shared regulatory modules across excitable cell types, particularly ion channel regulation and metabolic support.^50,51^ The findings are exploratory but hints on linked biology of early period of MS severity.

The MR analyses did not support causal associations of 25-hydroxyvitamin D, BMI, consistent with prior evidence that these factors failing to exert modulatory effects on disability progression.^37,38^ The lifetime smoking index also did not exhibit any causal relationship. Smoking has been associated with faster progression of the disease,^39^ but the effect is reported to diminish after quitting smoking.^40^ Since this study measured the first recorded EDSS scores after diagnosis, it is plausible that pwMS had significant motivation to change their smoking behavior following diagnosis, as a result weakening the causal relationship. Educational attainment suggested a nominal bidirectional causal association with early MS severity. This bidirectional outputs align with the broader concept of cognitive reserve and resilience, in which higher educational attainment may buffer against cognitive damage.^41,42^ Conversely, genetic variants that accelerate early disease severity may influence neural integrity in prodromal and early phase of MS during adolescence and early adulthood, leading to cognitive difficulties and having fewer years of education.^43^

In silico drug-prioritization analyses nominated compounds targeting the GWAS-prioritized genes *TNFSF4*, *TNFSF18*, and *ADAMTS2*. Despite the drug-gene link are more of hypothesis-generating, the findings narrow down the candidate drug and therapeutic components that could be informative in further validation experiments to establishing disease modifying treatment in MS.

This study has limitations. First, all participants were of European genetic ancestry, limiting generalizability to other ancestries. Second, relapse status at the time of EDSS assessment was unavailable in the Italian cohort during the data analyses phase. However, EDSS measurements were clinically guided to be conducted outside periods of recent relapse. Approximately 1.7% of the EDSS measurements (discovery: 116/5593, replication: 4/1616) may have been used in the late MS severity GWAS, since the individuals (N = 120) had only one measurement recorded in the registries. However, the minimal overlap suggests a negligible impact on our findings. Finally, in MR analyses, instrumental variable assumption violation cannot be fully excluded, despite our sensitivity MR analyses displayed no sign of horizontal pleiotropy.

In conclusion, our study provides insight into the genetic architecture of early MS severity and nominates effector biology for functional follow-ups. Further experimental work is needed to clarify the mechanism and evaluate therapeutic relevance for the early stages of MS.

## Data Availability

All available data in the present study are in the manuscript.

## Acknowledgments

We would like to thank Ingileif Jónsdóttir and Kári Stefánsson at Amgen deCODE Genetics and University of Iceland, Reykjavik, Iceland, for genotyping the Swedish cohorts and providing genetic expertise. We thank all study participants, clinical staff and laboratory technicians at involved centers.

## Author Contributions

PS, IK, and SJ conceived of and designed the study. KS, MS, AM, LA, JH, TO, FP, FC, MR, MF, FE, and PS collected data. SJ, FC, AH, and PS performed genotyping and/or quality control. SJ, FC analysed the data. PS, IK, and KS supervised this study. SJ, KS, PS and IK drafted the manuscript. MS, AH, AM, LA, JH, MJ, TO, FP, FC, MR, MF, and FE revised and edited the manuscript.

## References

1. the International Multiple Sclerosis Genetics Consortium. Class II HLA interactions modulate genetic risk for multiple sclerosis. Nat Genet. 2015;47(10):1107–1113. doi:10.1038/ng.3395

2. International Multiple Sclerosis Genetics Consortium, Patsopoulos NA, Baranzini SE, et al. Multiple sclerosis genomic map implicates peripheral immune cells and microglia in susceptibility. Science. 2019;365(6460):eaav7188. doi:10.1126/science.aav7188

3. International Multiple Sclerosis Genetics Consortium, Harroud A, Stridh P, et al. Locus for severity implicates CNS resilience in progression of multiple sclerosis. Nature. 2023;619(7969):323–331. doi:10.1038/s41586-023-06250-x

4. Sahi N, Haider L, Chung K, et al. Evaluating multiple sclerosis severity loci 30 years after a clinically isolated syndrome. Brain Communications. 2024;6(6). doi:10.1093/braincomms/fcae443

5. Campagna MP, Havrdová E, Horáková D, et al. No evidence for association between rs10191329 severity locus and longitudinal disease severity in 1813 relapse-onset multiple sclerosis patients from the MSBase registry. Multiple Sclerosis Journal. 2024;30(9):1216–1220. doi:10.1177/13524585241240406

6. Stridh P, Huang J, Kockum I. From discovery to replication: Power and definitions matter for multiple sclerosis severity. Multiple Sclerosis Journal. 2024;30(10):1385–1386. doi:10.1177/13524585241265021

7. King EA, Davis JW, Degner JF. Are drug targets with genetic support twice as likely to be approved? Revised estimates of the impact of genetic support for drug mechanisms on the probability of drug approval. doi:10.1371/journal.pgen.1008489

8. Chang C, Chow CC, Tellier LC, Vattikuti S, Purcell S, Lee JJ. Second-generation PLINK: rising to the challenge of larger and richer datasets. GigaScience. 2015;4(1). doi:10.1186/s13742-015-0047-8

9. Kurtzke JF. Rating neurologic impairment in multiple sclerosis. Neurology. 1983;33(11):1444–1444. doi:10.1212/wnl.33.11.1444

10. Aschard H, Vilhjálmsson BJ, Joshi AD, Price AL, Kraft P. Adjusting for Heritable Covariates Can Bias Effect Estimates in Genome-Wide Association Studies. The American Journal of Human Genetics. 2015;96(2):329–339. doi:10.1016/j.ajhg.2014.12.021

11. Bulik-Sullivan B, Loh PR, Finucane HK, et al. LD Score regression distinguishes confounding from polygenicity in genome-wide association studies. Nature Genetics. 2015;47(3). doi:10.1038/ng.3211

12. He Y, Koido M, Shimmori Y, Kamatani Y. GWASLab: a Python package for processing and visualizing GWAS summary statistics | Jxiv, JST □□□□□□□□□. Accessed October 13, 2025. https://jxiv.jst.go.jp/index.php/jxiv/preprint/view/370

13. Willer CJ, Li Y, Abecasis GR. METAL: fast and efficient meta-analysis of genomewide association scans. Bioinformatics. 2010;26(17). doi:10.1093/bioinformatics/btq340

14. Lee S, Yang J, Goddard ME, Visscher PM, Wray NR. Estimation of pleiotropy between complex diseases using single-nucleotide polymorphism-derived genomic relationships and restricted maximum likelihood. Bioinformatics. 2012;28(19). doi:10.1093/bioinformatics/bts474

15. Yang J, Lee SH, Goddard ME, Visscher PM. GCTA: A Tool for Genome-wide Complex Trait Analysis. Am J Hum Genet. 2011;88(1):76–82. doi:10.1016/j.ajhg.2010.11.011

16. Evans LM, Tahmasbi R, Vrieze S, et al. Comparison of methods that use whole genome data to estimate the heritability and genetic architecture of complex traits. Nature Genetics. 2018;50(5). doi:10.1038/s41588-018-0108-x

17. Finucane HK, Reshef Y, Anttila V, et al. Heritability enrichment of specifically expressed genes identifies disease-relevant tissues and cell types. Nature Genetics. 2018;50(4). doi:10.1038/s41588-018-0081-4

18. Zou Y, Carbonetto P, Wang G, Stephens M. Fine-mapping from summary data with the “Sum of Single Effects” model. doi:10.1371/journal.pgen.1010299

19. Benner C, Spencer CCA, Havulinna AS, Salomaa V, Ripatti S, Pirinen M. FINEMAP: efficient variable selection using summary data from genome-wide association studies. Bioinformatics. 2016;32(10):1493–1501. doi:10.1093/bioinformatics/btw018

20. Hof JP, Speed D. LDAK-KVIK performs fast and powerful mixed-model association analysis of quantitative and binary phenotypes. Nature Genetics. 2025;57(9):2116–2123. doi:10.1038/s41588-025-02286-z

21. Speed D, Holmes J, Balding DJ. Evaluating and improving heritability models using summary statistics. Nature Genetics. 2020;52(4):458–462. doi:10.1038/s41588-020-0600-y

22. De Leeuw CA, Mooij JM, Heskes T, Posthuma D. MAGMA: Generalized Gene-Set Analysis of GWAS Data. Tang H, ed. PLoS Comput Biol. 2015;11(4):e1004219. doi:10.1371/journal.pcbi.1004219

23. Manouchehrinia A, Westerlind H, Kingwell E, et al. Age Related Multiple Sclerosis Severity Score: Disability ranked by age. Mult Scler. 2017;23(14):1938–1946. doi:10.1177/1352458517690618

24. Lublin FD, Häring DA, Ganjgahi H, et al. How patients with multiple sclerosis acquire disability. Brain. 2022;145(9):3147–3161. doi:10.1093/brain/awac016

25. Manian M, Motallebnezhad M, Nedaeinia R, et al. Comparison of OX40 expression in patients with multiple sclerosis and neuromyelitis optica as an approach to diagnosis. *Allergy*, Asthma & Clinical Immunology. 2023;19(1):19. doi:10.1186/s13223-023-00772-9

26. Huseby ES, Kamimura D, Arima Y, Parello CS, Sasaki K, Murakami M. Frontiers | Role of T cell —glial cell interactions in creating and amplifying central nervous system inflammation and multiple sclerosis disease symptoms. Accessed December 8, 2025. https://www.frontiersin.org/journals/cellular-neuroscience/articles/10.3389/fncel.2015.00295/full

27. Mousavi SF, Soroosh P, Takahashi T, et al. OX40 Costimulatory Signals Potentiate the Memory Commitment of Effector CD8+ T Cells. J Immunol. 2008;181(9):5990–6001. doi:10.4049/jimmunol.181.9.5990

28. Ray A, Basu S, Williams CB, Salzman NH, Dittel BN. A Novel IL-10–Independent Regulatory Role for B Cells in Suppressing Autoimmunity by Maintenance of Regulatory T Cells via GITR Ligand. J Immunol. 2012;188(7):3188–3198. doi:10.4049/jimmunol.1103354

29. Hwang H, Lee S, Lee WH, Lee HJ, Suk K. Stimulation of glucocorticoid-induced tumor necrosis factor receptor family-related protein ligand (GITRL) induces inflammatory activation of microglia in culture. Journal of Neuroscience Research. 2010;88(10):2188–2196. doi:10.1002/jnr.22378

30. Lemarchant S, Pruvost M, Montaner J, et al. ADAMTS proteoglycanases in the physiological and pathological central nervous system. Journal of Neuroinflammation. 2013;10(1):899. doi:10.1186/1742-2094-10-133

31. Cross AK, Haddock G, Surr J, et al. Differential expression of ADAMTS-1, -4, -5 and TIMP-3 in rat spinal cord at different stages of acute experimental autoimmune encephalomyelitis. Journal of Autoimmunity. 2006;26(1):16–23. doi:10.1016/j.jaut.2005.09.026

32. Haddock G, Cross AK, Plumb J, et al. Expression of ADAMTS-1, -4, -5 and TIMP-3 in normal and multiple sclerosis CNS white matter. Mult Scler. 2006;12(4):386–396. doi:10.1191/135248506ms1300oa

33. Bulik-Sullivan B, Finucane HK, Anttila V, et al. An atlas of genetic correlations across human diseases and traits. Nature Genetics. 2015;47(11):1236–1241. doi:10.1038/ng.3406

34. Tiane A, Schepers M, Reijnders RA, et al. From methylation to myelination: epigenomic and transcriptomic profiling of chronic inactive demyelinated multiple sclerosis lesions. Acta Neuropathol. 2023;146(2):283–299. doi:10.1007/s00401-023-02596-8

35. Humphries ESA, Dart C. Neuronal and Cardiovascular Potassium Channels as Therapeutic Drug Targets. J Biomol Screen. 2015;20(9):1055–1073. doi:10.1177/1087057115601677

36. Skavinska O, Rossokha Z, Fishchuk L, Gorovenko N. *RFC* and *VDR* -mediated genetic regulation of brain folate transport in patients with multiple sclerosis. Human Gene. 2025;44:201399. doi:10.1016/j.humgen.2025.201399

37. Mahler JV, Solti M, Apóstolos-Pereira SL, Adoni T, Silva GD, Callegaro D. Vitamin D3 as an add-on treatment for multiple sclerosis: A systematic review and meta-analysis of randomized controlled trials. Multiple Sclerosis and Related Disorders. 2024;82:105433. doi:10.1016/j.msard.2024.105433

38. Manuel Escobar J, Cortese M, Edan G, et al. Body mass index as a predictor of MS activity and progression among participants in BENEFIT. Mult Scler. 2022;28(8):1277–1285. doi:10.1177/13524585211061861

39. Healy BC, Ali E, Guttmann CRG, et al. Smoking and Disease Progression in Multiple Sclerosis. Arch Neurol. 2009;66(7):858–864. doi:10.1001/archneurol.2009.122

40. Rodgers J, Friede T, Vonberg FW, et al. The impact of smoking cessation on multiple sclerosis disease progression. Brain. 2022;145(4):1368–1378. doi:10.1093/brain/awab385

41. Liu Y, Lu G, Liu L, He Y, Gong W. Frontiers | Cognitive reserve over the life course and risk of dementia: a systematic review and meta-analysis. doi:10.3389/fnagi.2024.1358992

42. Seyedsalehi A, Warrier V, Bethlehem RAI, Perry BI, Burgess S, Murray GK. Educational attainment, structural brain reserve and Alzheimer’s disease: a Mendelian randomization analysis. Brain. 2022;146(5):2059–2074. doi:10.1093/brain/awac392

43. Harding KE, Kreft KL, Ben-Shlomo Y, Robertson NP. Prodromal multiple sclerosis: considerations and future utility. J Neurol. 2024;271(4):2129–2140. doi:10.1007/s00415-023-12173-4

