## Supplementary for "Genome-Wide Study of Early Disability Severity in Multiple Sclerosis"

### eSupplementary Text

#### Standard Protocol Approvals, Registrations, and Patient Consents

For the discovery cohort, 18119 participants were recruited from four Swedish nationwide epidemiological studies: EIMS,[1] GEMS,[2] IMSE,[3] COMBAT-MS,[4] and one clinical STOP-MS study.[5] Clinical phenotypes were obtained through linkage to the Swedish multiple sclerosis (MS) registry.[6] In addition, harmonized genetic data and lifestyle questionnaires were available. For the replication cohort, 1891 people with MS (pwMS) were recruited at the MS Center of San Raffaele Hospital (Milan, Italy) from 2003 to 2022, and 517 pwMS from the MultipleMS (MMS) prospective cohort (2017-2022), which is composed of treatment-naive pwMS across seven European countries (Sweden, Italy, Germany, Belgium, Spain, Denmark, and Norway).[7] Written informed consent was obtained from all participants, and the studies were approved by the local ethics committee.  MS Individuals were diagnosed by neurologists based on Mcdonald criteria[8] and those with at least one disability measure were included. Neurological disability was assessed using the Expanded Disability Status Scale (EDSS)[9], a measurement based on impairments in eight functional systems, with each system representing a neuronal network responsible for a particular task. In the Swedish cohorts, we excluded individuals lacking genetic data, without MS or disease-status information, with disease courses other than progressive or bout-onset MS, or with a negative interval from disease onset to the first EDSS assessment (n=5,733). In the Italian cohort, we excluded pwMS who lacked genetic data or had a disease duration of zero or less at the first EDSS assessment (n=585). In the MMS cohort, we excluded pwMS without genetic data (n=54). Because EDSS variability is known to be higher during the early stages of MS, relapse-free EDSS scores, defined as assessments without a relapse in the preceding 30 days, were used for the discovery and MultipleMS cohorts (n=6,990), as recommended by Ramotello et al.[10] The relapse information was absent in the Italian cohort in the time of analyses. We also employed the EDSS measurement that had disease duration ≤ 18 years since the late MS severity genome-wide association study (GWAS) had mean year of 18.2 years for their disease duration, ensuring our EDSS measurements are of earlier period. The pwMS that satisfied the disease duration cutoff and complete information for the regression were 5593 for Swedish, 1160 for Italians, and 456 for MultipleMS cohorts. These initial EDSS score for each participant was transformed into an Age-Related Multiple Sclerosis Severity (ARMSS) score.[11] Descriptive statistics of the discovery and replication cohorts are in Table 1.

#### Genotyping, quality control, and imputation

For Swedish and MMS cohorts, DNA was extracted from the whole blood. Most of the samples, this was done at KI biobank Core Facility, Stockholm, Sweden. using Chemagen kits based on magnetic bead separation and some were extracted at Center of Molecular Medicine, Stockholm, Sweden, for these we used Qiagen kits. DNA was stored at -20C after extraction. The genotyping was performed using illumina Global Screening Arrays (GSA, GSA2) and Human Omni Express (OE) at deCODE Genetics in Iceland for the Swedish cohorts and at FIMM Genomics Services for MMS. Sample- and variant-level quality control (QC) and imputation for GSA, OE, and MMS cohorts have been described in detail in the late MS severity GWAS.[12] For GSA2, we removed mitochondrial DNA, duplicates, and multi-allelic variants, and lifted the genome build from GRCh38 to GRCh37 using triple-liftover. Triple-liftover is a tool that allows SNPs in regions inverted between the genome builds to be correctly converted, which imputation servers such as TOPMed quality control pipelines fail to do.[13] Markers were removed if they had >5% missingness, differential Hardy-Weinberg equilibrium (HWE) in controls (P-value thresholds: $P < 1 \times 10^{-6}$ for MAF > 0.1, $P < 1 \times 10^{-5}$ for 0.05 < MAF <= 0.1, and $P < 1 \times 10^{-4}$ for 0.01 <= MAF <= 0.05), differential missingness between cases and controls ($P < 1 \times 10^{-4}$), or minor allele frequency (MAF) < 0.01. For sample QC, we excluded samples with >5% missingness, heterozygosity outliers defined by an inbreeding coefficient $|F| < 0.05$, and relatedness at third degree or closer (using PLINK 2.0,[14] KING cutoff = 0.0442). Population structure was assessed via principal component analysis (PCA) on both GSA2 and the 1000 Genomes European (1KG EUR; EUR = European) cohort using hard-called linkage disequilibrium (LD)-pruned genotypes via the bigsnpr R package (v1.12.18). Outliers were removed using a modified probabilistic local outlier factor statistic, which better captures outliers than the traditional six standard deviations from the mean PC scores.[15] Phasing was conducted with Eagle2[16] using the Haplotype Reference Consortium (HRC) panel (EUR GRCh37/hg19, release 1.1).[17] The genome was segmented into 20Mb chunks with an additional 5Mb overlap in flanking regions. Imputation was performed using Minimac4 (v1.0.2)[18] in overlapping 500kb windows to maintain LD continuity. Imputed chunks were merged to produce a single VCF file per chromosome. Imputed genotypes were evaluated per chromosome and chip to ensure continuity, consistency in imputation score ($R^2$), and alternate allele frequency differences with the reference panel (eFigure 2). Mean imputation scores and total frequencies from Minimac4 were analyzed across various MAF bins (eFigures 3-4). Based on these analyses, post-imputation QC retained variants with MAF > 0.01 and imputation $R^2 > 0.3$.

For Italians (OSR), genomic DNA was extracted from whole blood samples using standard phenol-chloroform procedures or commercially available extraction kits (QIAamp DNA Blood Kits, Qiagen, Hilden, Germany; Maxwell DNA Purification System, Promega, Madison, WI, USA), according to the manufacturers' protocols. DNA samples were stored at -20°C until analysis. Genome-wide genotyping was performed using Illumina® Human660W-Quad, HumanOmniExpress-12 BeadChip, HumanOmniExpress-24 BeadChip, HumanOmni2.5 BeadChip, or Global Screening Array platforms. Genotyping was carried out at the IRCCS San Raffaele Scientific Institute, except for samples genotyped with the Human660W-Quad platform, which were processed within the WTCCC2 project.[19] Imputation was performed against the Haplotype Reference Consortium Rel 1.1 2016 EUR on the GRCh37/hg19 genome as a reference panel, using Eagle v2.4 for the pre-phasing of haplotypes and Minimac4 for the imputation of the missing genotypes. PwMS genotyped on the Quad-660, HumanOmniExpress/HumanOmni-2.5, and Global Screening Array platforms were imputed individually. Imputed SNPs with $R^2 < 0.6$ and/or MAF < 0.01 in the merged cohorts were excluded from the analysis. Standard per-sample and per-single-nucleotide polymorphism (SNP) quality controls were implemented using PLINK 1.9.[20] Samples with sex mismatch, genotype call-rate below 95%, high heterozygosity rate, high relatedness and ancestry outliers according to multi-dimensional scaling analysis were excluded; The variants with MAF < 1%, genotypic call-rate < 97% or departing from for HWE at $P < 1 \times 10^{-4}$ were excluded.

All cohort variants were reported using rsIDs and GRCh37/hg19 chromosome:position:reference:alternate coordinates in accordance with the hg19 reference panel.

#### Fine-mapping for causal variants identification

We used two methods for fine-mapping for robustness: SuSiE-RSS[21] and FINEMAP.[22] SuSiE-RSS λ was near zero and kriging diagnostics supported overall concordance between the LD matrix and association z scores in both loci (rs929769 $\lambda = 1.69 \times 10^{-6}$; rs1234317 $\lambda = 2.53 \times 10^{-6}$). At chromosome 5, fine-mapping formed a single-variant 95% credible set (5:178727997:T:C, rs929769) in SuSiE (posterior inclusion probability (PIP) = 0.964) and a single causal variant model in FINEMAP with the highest posterior probability (PP), 0.643 (eFigure 5A,C). SNP-level PIP was highly concordant between FINEMAP and SuSiE-RSS (Pearson r = 1.00, Spearman rho = 0.938; eFigure 5E, eTable 5). At chromosome 1, SuSiE-RSS yielded two variants, 1:173133986:A:G (rs6657136, PIP=0.561) and 1:173187775:C:T (rs1234317, PIP=0.399), for the primary 95% credible set, but the secondary credible set had diffuse effects composed of 1339 SNPs (eFigure 5B, D). FINEMAP had higher probability of two-causal-SNP model (PP = 0.620) over single causal-SNP model (PP=0.374), but the highest two-SNP configuration had extremely low PP (=0.0090). Across configurations, SNP-level FINEMAP PP was the highest for rs6657136 (PP=0.527), followed by rs1234317 (PP=0.442). SNP-level PIP was concordant between FINEMAP and SuSiE-RSS in chromosome 1 (Pearson r=0.997, Spearman rho=0.982; eFigure 5F, eTable 6). After conducting the fine-mapping analyses, we prioritized three most supported candidate causal SNPs for downstream analyses: rs929769, rs1234317, and rs6657136.

#### rs1234317, rs6657136, and rs929769 associations with MS time-to-event phenotypes

We identified a subset of pwMS from the discovery cohort with a minimum of three clinical assessments at intervals of at least six months, excluding those assessments within 30 days after any relapses. This resulted in 60,781 study visits from 7,105 pwMS. Three time-to-event traits were applied: rate of EDSS progression, time to clinically meaningful worsening,[23] and time to EDSS 6. Year of birth, sex, age at onset, genotyping chip, imputation wave, and principal components were adjusted in the models. To examine the effect of SNPs on the rate of disability progression, a generalized linear mixed model (GLMM) was used. The primary variable of interest was the interaction between genotype and time within the study, with individuals treated as random effects. This study utilized penalized quasi-likelihood estimation, as executed through the glmmPQL function from the MASS R package (version 7.3.65), to address the non-normal distribution of EDSS scores. To examine the effect of the SNPs on the time to clinically meaningful worsening, we used the following definition; an increase in EDSS by 1.0, if the baseline was < 5.5, and by 0.5 if the baseline was >= 5.5.[23] To enhance specificity, the EDSS increase had to be maintained at subsequent visits and for at least 24 weeks. Cox proportional hazard was calculated with the Survival package in R (version 3.8.3), and the proportional hazards assumption was tested via Schoenfeld residuals and satisfied by either adding an interaction term or allowing the baseline hazard function to differ between categorical variables. Finally, we performed survival analyses for the time to reach EDSS 6, a critical disability milestone in MS progression. Given the discrete nature of the EDSS assessments, interval censoring was implemented to account for the unobserved exact time to reach an EDSS score of 6.0. Left censoring was also considered for those who had already reached this threshold before the first EDSS measurement, and right censoring was applied to those who had not reached an EDSS score of 6.0 by the end of their assessments. Cox proportional hazards with the applied censorings were analyzed using the icenReg package (version 2.0.16) in R. The time to sustained EDSS 6.0 were analyzed as sensitivity analyses of time to EDSS 6.0, which requires all subsequent scores after reaching EDSS 6.0 to remain or be above 6.0. The same censorings and R package was used for estimation. For all the time-to-event analyses, hazard ratios (HR) were calculated using hard-called dosages, and P-values less than 0.0083 (N=6) were considered significant after Bonferroni correction.

We observed that the carriers of rs1234317_T had modestly higher EDSS scores over the disease duration ($\beta$ = 0.0980, SE = 0.0429, P = 0.0226) without evidence of an altered rate of EDSS progression (P = 0.442, eFigure 7A), a significantly shorter time to EDSS 6 (hazard ratio [HR] = 1.15, SE = 0.0504, P = 0.00688, eFigure 7B), and a nominally significant association with time to sustained EDSS 6 (HR = 1.12, SE = 0.0532, P = 0.0369, eFigure 7C). The time to 24-weeks confirmed disability worsening did not achieve statistical significance but exhibited a consistent trend in direction of effect (HR = 1.05, SE = 0.0528, P = 0.312, eFigure 7D). For rs6657136_G, the carriers had modestly lower EDSS scores over time with nominal significance ($\beta$ = -0.0887, SE = 0.0405, P = 0.0285) without evidence of altered rate of EDSS progression (P=0.411; eFigure 8A), a nominally significantly longer time to EDSS 6 (HR = 0.91, SE = 0.0452, P = 0.0367; eFigure 8B), non-significant associations with time to sustained EDSS 6 (HR = 0.927, SE = 0.0519, P = 0.146; eFigure 8C) and time to 24-weeks confirmed disability worsening (HR = 1.01, SE = 0.0512, P = 0.879, eFigure 8D). For rs929769_C, carriers had significantly higher EDSS scores over time ($\beta$ = 0.159, SE = 0.0472, P = 0.0007) without evidence of altered rate of EDSS progression (P=0.316; eFigure 9A), a non-significant association yet consistently shorter trend of time to EDSS 6 (HR = 1.09, SE = 0.0540, P = 0.0947; eFigure 9B), time to sustained EDSS 6 (HR = 1.07, SE = 0.0609, P = 0.262; eFigure 9C), and 24-week confirmed disability worsening (HR = 1.01, SE = 0.0367, P = 0.753; eFigure 9D).

##### Multi-omics integrated gene prioritization and annotation

To prioritize candidate effector genes, we applied an integrative SNP-to-gene framework to the lead variants and their LD proxies from the 1000 Genomes phase 3 European reference panel ($r^2$ >= 0.6), with build-aware variant harmonization using Ensembl and UCSC liftOver.[24–26] Evidence sources included: (1) ANNOVAR/refGene positional annotation of lead variants and LD proxies;[27] (2) molecular quantitative trait locus (QTL) evidence from GTEx v8 and eQTL Catalogue expression QTL (eQTL)/splicing QTL (sQTL) resources;[28–30] (3) chromatin/regulatory evidence from Hi-C, ABC enhancer-gene maps, and promoter-capture Hi-C (pcHi-C);[31–33] (4) precomputed Open Targets Platform/Genetics L2G predictions for credible sets matching lead or proxy variants, using L2G >= 0.5 as the threshold;[34–37] (5) precomputed cS2G scores, using cS2G >= 0.5;[38,39] (6) post hoc FUSION transcriptome-wide association study (TWAS) with CommonMind Consortium dorsolateral prefrontal cortex expression weights;[40,41] and (7) coloc-ABF and coloc-SuSiE posterior evidence.[42,43] Prespecified base weights from the publicly available database were positional=1.0, eQTL=2.0, sQTL=1.5, cS2G=2.5, L2G=2.5, Hi-C=1.0, ABC=2.0, PCHi-C=1.5. The weighting scheme is literature-prior evidence hierarchy: proximity and generic contact evidence were retained as baseline support, direct QTL[28–30] and validated enhancer-promoter models[32,33] were weighted more strongly, supervised composite SNP-to-gene models were weighted highest among base methods,[34,38] TWAS had moderate evidence weight because LD and correlated predicted expression can confound gene-level associations, and colocalization had stronger weights than TWAS because PP4 estimates whether the GWAS and molecular QTL signals share a causal variant. When 0.5 <= PP4 < 0.75, 2.0 were given, and in 0.75 <= PP4, 3.0 was granted as the weight.[41–45] The complete weighted scores were computed as:

$$weighted\ \ score\ =\ \Sigma_i\ \ w_i \cdot I_i$$

where $I_i = 1$ when evidence exists and $I_i = 0$ otherwise. The downstream annotation of the top prioritized genes for each SNP signal used GTEx Analysis Release V10 and Human Protein Atlas single-cell/single-nucleus resources.[28,46,47]

The LD expansion identified 23 proxy SNPs for the three candidate causal SNPs and we prioritized the top candidate effectors that are supported by multiple classes of evidence as mentioned. For rs1234317, *TNFSF4* outweighed other candidates in the weighted score (6.0), with cS2G score reaching maximum score 1.0, L2G score 0.863, and positionally the closest. *TNFSF4* is expressed in the brain cerebellum, immune tissues, Epstein-Barr virus (EBV)-transformed lymphocytes, and cultured fibroblasts (eFigure 10A). In cell types, strong expression occurred in astrocytes with moderate neuronal and oligodendroglia expression, as well as in natural killer (NK) cells, T cells, and B cells (eFigures 11A, 12A).

For rs6657136, *TNFSF18* was the strongest scored gene (5.5), evidenced by positional mapping, eQTL, Hi-C, and pcHi-C. In the eQTL Catalogue, the signal was associated with *TNFSF18* expression in dorsolateral prefrontal cortex ($P = 2.21 \times 10^{-11}$), and Hi-C and pcHi-C associated the variant regions to *TNFSF18* promoters across multiple tissues and cell types. It is notably expressed in the various brain tissues and liver portal tract (eFigure 10B). In cell types, strong expressions were detected in basal squamous and squamous epithelial cells with moderate expressions in macrophages, monocytes, Muller glia cells, excitatory neurons, and oligodendroglia (eFigure 11B). Within the brain, expression is in various neuronal, CNS macrophage, and glial cells (eFigure 12B).

For rs929769 locus, *ADAMTS2* outscored the other genes as an effector (3.5), grounded by positional mapping and cS2G, with cS2G score 1.0. In tissues and cell-types, *ADAMTS2* is reported to be highly expressed in fibroblasts, but also expressed in macrophage and oligodendroglial cells (eFigures 10C, 11C). In single-nucleus RNA-seq datasets, we observed the enrichments in neuronal clusters (eFigure 12C).

#### Gene-level association and pathway analyses

We performed a genome-wide gene association scan (GWGAS) using two complementary approaches. First, we implemented MAGMA v1.10[48] in multi-model mode using pooled individual-level genotype data from discovery and replication cohorts. Multi-model mode of MAGMA aggregates SNP-level statistics into gene-level P-values combining the results of the three base models (Principal Components Regression, SNP-wise Mean, and SNP-wise Top), which gives the most even distribution of power and sensitivity for a broader range of various genetic architectures. Gene-set analysis was subsequently performed using competitive framework, accounting for gene size, gene density, and LD between genes. To explore biological relevance, we selected genes with nominal significance ($P < 0.05$) present in the Ingenuity Knowledge Base and subjected them to canonical pathway, upstream regulator, and toxicity analyses. Enrichment significance was assessed using right-tailed Fisher's exact tests with Benjamini-Hochberg correction. Second, we applied LDAK-KVIK-GBAT, a mixed-model method that combines LDAK-KVIK[49] with LDAK-GBAT,[50] which has been shown to detect substantially more significant genes than MAGMA for quantitative phenotypes. Gene-level results from LDAK-KVIK-GBAT were further analyzed in FUMA to identify tissues in which mapped genes are differentially expressed (Bonferroni-corrected hypergeometric test) and enriched in gene sets from MSigDB, WikiPathways, and KEGG.

The linear model (MAGMA) did not identify any gene-level association, but the mixed model (LDAK-KVIK-GBAT) identified 62 genes surpassing the Bonferroni threshold and 189 genes at false discovery rate (FDR) significance (eTables 10-11, eFigure 13). Using the 189 mapped genes, we examined tissue- and cell-type-specific expression patterns. In 54 GTEx v8 tissue-specific differentially expressed gene (DEG) sets, we observed significant overlap between the genes and downregulated DEGs in several brain subregions (anterior cingulate cortex, cerebellum, hippocampus, nucleus accumbens, and putamen), upregulated DEGs in EBV-transformed lymphocytes, and bidirectionally regulated DEGs in the left ventricle (eTable 12, eFigure 14). In the 30 general tissue DEG sets, we observed significant overlap with downregulated DEGs in brain tissues (eTable 13, eFigure 15). The brain downregulation pattern may indicate shared transcriptional characteristics with reduced expression in brain tissues, while the lymphocyte signal aligns with immune activation. The cardiac tissue signal may reflect shared signatures of cellular energy metabolism or stress response across excitable cell types. Three pathway enrichment analyses were performed. Competitive gene-set analysis via MAGMA did not produce significant pathways after correction (eTable 14, eFigure 16A), nor did IPA canonical pathway analysis (eTable 15, eFigure 16B). FUMA highlighted five gene ontologies: sinoatrial node cell development and differentiation, cardiac pacemaker cell differentiation, folic acid transport, and folic acid binding (eTable 16, eFigure 16C-D). Although non-canonical MS pathways, these may reflect shared regulatory modules across excitable cell types, particularly ion channel regulation and metabolic support.[51,52] Folic acid metabolism is crucial for methylation, DNA stability, nucleotide metabolism, and remyelination.[53,54] Upstream regulator analysis identified 19 significant molecules ($q < 0.05$), including STING1 (cGAS-STING interferon axis), IGFBP5 (growth factor binding), and PARP3 (DNA repair and chromatin remodeling), implicating the convergence of innate immune activation, DNA-damage response, and ferroptosis-mediated neurodegeneration (eTable 17).[55–57] These pathways affect neuronal survival, oligodendrocyte differentiation, and microglial activation during demyelination-remyelination cycles.[55,57,58] Toxicity analysis did not reveal significant associations (eTable 18). Collectively, the gene-level and pathway analyses point to enrichment of early MS severity genes in immune, DNA repair, and metabolic pathways that converge on central nervous system (CNS) integrity.

#### Genetic correlations of early MS severity with MS susceptibility, late MS severity, and non-MS traits

To verify the genetic relationship between early MS severity and MS susceptibility, we obtained 208 (196 non-major histocompatibility complex [MHC], 12 MHC) variants selected from 232 autosomal MS susceptibility associations of the IMSGC susceptibility GWAS[59] by excluding variants that were palindromic and with P > 5 x $10^{-8}$, absent in our dataset after QC. To examine MHC associations, we exclusively considered non-palindromic SNPs and utilized rs3135388 as a surrogate marker for the *human leukocyte antigen (HLA)-DRB1**15:01 allele. The association of these variants with early MS severity was evaluated using linear regression framework. The results were subsequently meta-analyzed across the European cohorts. A Bonferroni correction was applied to account for multiple testing (P = 2.40 x $10^{-4}$). We also tested the LD relationships with the two SNPs of our early MS severity GWAS using PLINK 2.0.[14] An exact two-sided binomial test was employed to evaluate the concordance in the direction of the effect of the SNPs between MS susceptibility and early MS severity. To assess the cumulative impact of MS susceptibility on disability outcomes, we developed polygenic scores (PGS) with respect to the selected MS susceptibility variants and regressed the first ARMSS scores on that PGS in the GSA2 chip cohort. Swedish GSA2 chip cohort was selected since the participants are not expected to overlap with those of MS susceptibility GWAS,[59] removing the possibility of the inflation of the association statistic.[60] We implemented in two ways: first regressing on the PGS adjusted for the year of birth, sex, and the principal components, and second regressing the first ARMSS scores without the PGS. We then calculated the $R^2$ difference of the two and reported as the incremental $R^2$.

We examined the association of rs10191329 and rs149097173, the SNPs of late MS severity GWAS[12] with early MS severity using the same covariates for each genotyping chip. rs10191329 was tested in the meta-analyzed cohort, but rs149097173 was tested in the discovery cohort due to data availability. We also tested the LD relationships of these two SNPs with the two SNPs of our early MS severity GWAS using PLINK 2.0. To assess the cumulative impact of late MS severity on early disability outcomes, we developed a polygenic score with respect to suggestively significant late MS severity GWAS SNPs, which the threshold defined by their study (P < 5 x $10^{-6}$), retained after LD clumping ($r^2 < 0.01$), minor allele frequency >= 0.01, and imputation score > 0.8. We then calculated the incremental $R^2$ in the same way as for susceptibility and early MS severity.

For genetic comparison with other non-MS traits, we evaluated phenome-wide associations for our SNPs of interest using Open Targets Genetics,[35] which combines the GWAS Catalog,[61] UK Biobank,[62] and FinnGen.[63] We did not expect any sample overlap with any of the traits, thus LDSC intercept was constrained on no shared population stratification. Benjamini-Hochberg adjustment was made for multiple testing correction.

Following are the results. Heritability enrichment analyses of specifically expressed gene sets showed distinct immune-cell enrichment for MS susceptibility and CNS-cell enrichment for late MS severity, whereas early MS severity showed no significant immune- or CNS-cell enrichment (Figure 2; eTable 3).

For the overlap between the early MS severity and susceptibility, one MS susceptibility SNP, rs4409785, was associated with early MS severity after Bonferroni correction ($P$=0.000145) and 17 SNPs were nominally associated ($P$<0.05; eTable 19). One nominally associated SNP (rs7454108) maps to the HLA region, but the SNP with strongest LD to DRB1*15:01, rs3135388, is not associated with early MS severity ($P$ = 0.3374). The proportion of susceptibility variants demonstrating a concordant direction of effect with the early MS severity GWAS variants significantly differed from what is expected by chance ($P$ = 0.0266; eTable 20). However, the LD between the early MS severity SNPs and the significant MS susceptibility SNPs were very low ($r^2 \leq 0.00133$, eTable 21, eFigure 17A) and the regression of early MS severity on the PGS with respect to MS susceptibility significant SNPs[59], revealed low incremental $R^2$ ($\Delta R^2$ = 0.00343) and non-significant association of PGS ($P$ = 0.146; eTable 22).

For the overlap between the early and late MS severity,[12] rs10191329, a genome-wide significant late MS severity SNP, was significantly associated with early MS severity ($\beta$ =0.0623, $P$ = 0.00588) in the same direction of effect, whereas rs149097173, a suggestive late MS severity SNP, was not ($\beta$ = -0.121, $P$ = 0.335; eTable 23). We additionally tested rs10191329 with early MS severity in non-overlapping cohorts with late MS severity GWAS participants[12] and found the SNP having the same direction of effect but not significant ($\beta$ =0.0327, $P$=0.546). The LD between the three SNPs of early MS severity and the late MS severity variants were low ($r^2 \leq 2.99 \times 10^{-4}$; eTable 24, eFigure 17B). The PGS with respect to suggestively significant SNPs of the late MS severity GWAS ($P = 5.0 \times 10^{-6}$) explained low variance of early MS severity ($\Delta R^2$ = 0.00211) and was not significantly associated with it ($P$ = 0.211; eTable 25).

A phenome-wide search across the GWAS Catalog, UK Biobank, and FinnGen for rs929769, rs6657136, and rs1234317 revealed no pleiotropic associations. In the genome-wide scale, there was a lack of evidence for shared genetic contributions between early MS severity and a range of neurological, psychiatric, cognitive, autoimmune, and aging phenotypes (eTable 26, eFigure 18).

#### Mendelian randomization

We applied Mendelian randomization (MR) analysis to investigate the effects of four exposures: 25-hydroxyvitamin D, BMI,[64,65] smoking,[66] and educational attainment,[66] which have been reported to be associated with MS susceptibility. Smoking and educational attainment were reported to be associated with late MS severity.[12] The GWAS used as proxies for exposure are listed in eTable 27. For each exposure, variants were selected based on two P-value thresholds ($P < 5 \times 10^{-8}$, $P < 1 \times 10^{-5}$), and LD was clumped ($r^2 < 0.001$) to ensure the independence of the variants. Palindromic variants were excluded from the study, and the included variants were evaluated for instrument strength (mean F-statistic > 10). The primary analysis used was inverse-variance-weighted (IVW) MR method with random effects. To assess heterogeneity among the genetic variants, which may suggest the presence of horizontal pleiotropy, we applied Cochran's Q-statistic and the MR pleiotropy residual sum and outlier global test (MR-PRESSO).[67,68] To further investigate the potential influence of horizontal pleiotropy, we employed four additional MR approaches: robust adjusted profile score (RAPS), weighted median, MR-PRESSO, and MR-Egger regression.[67] Consistent results across these methods can reduce the possibility of bias. While we reported beta coefficients for the MR-Egger regression, our primary focus was on the intercept as a test for unbalanced pleiotropy because the association estimate was underpowered. Analyses were conducted using the mr.raps 0.4.1, TwoSampleMR 0.6.10, MendelianRandomization 0.10.0, and MRPRESSO 1.0 R packages.

#### Genomics-based drug discovery

For the GWAS-prioritized genes, we used dual strategies. First, we utilized the Scalable Precision Medicine Open Knowledge Engine (SPOKE)[69] to identify approved/investigational compounds that 1) modulate the expressions of our GWAS-prioritized genes, *TNFSF4, TNFSF18,* and *ADAMTS2,* and bind their encoded proteins (ChEMBL,[70] BindingDB[71]). Complementary investigations were done in Therapeutic Target Database,[72] PharmGKB,[73] Open Targets Platform,[36] and DrugBank.[74] Second, acknowledging the limits of single-locus repurposing,[75] we applied TransPhar[56] to match compounds whose perturbational signatures inversely correlate with genetically regulated expression (GReX) patterns of MS severity genes predicted by FOCUS[76] across GTEx v7 tissues. We selected the top 10% of genes by Z score, then computed Spearman correlations between GReX profiles and L1000 compound signatures within CNS tissues/cell lines (MNEU.E, NEU, NPC, NPC.CAS9, NPC.TAK), yielding 82,680 tests. This was then expanded to additional GTEx tissues (N = 19) and L1000 cell types (N = 72). The discovery thresholds were FDR = 0.10 (per method guidance)[77] with rho < 0 to prioritize inverse correlations.

We report that *ADAMTS2* is associated with menadione, pirarubicin, dantrolene, epirubicin, neratinib, and imatinib. The protein product of *TNFSF4* was linked to magnesium sulfate heptahydrate, sodium sulfate, amlitelimab, oxelumab, and trebananib. TNFSF18 protein interacted with acetic acid (eTable 29). The nominated drugs are involved in immune modulation in T-cell activation and autoimmune disease propagation (oxelumab, amlitelimab[78]), BBB integrity, ameliorating neuroinflammation, and cerebral vasoconstriction (Dantrolene,[79] Neratinib,[80,81] Imatinib,[82] Magnesium sulfate heptahydrate, sodium sulfate[83]), reactive oxygen generation (Menadione,[84] Pirarubicin,[85,86] Epirubicin[86]), and short chain fatty acids (acetic acid).[87] The compounds that associates with the LDAK-KVIK-GBAT  FDR-significant genes are listed in eTable 30. Integration between the predicted genetically regulated expressions (GREx) of early MS severity and LINCS L1000 gene expression signature did not reveal any compounds with robust inverse correlation profile (eTable 31; eFigure 19).

#### eFigures

#### eFigure 1.

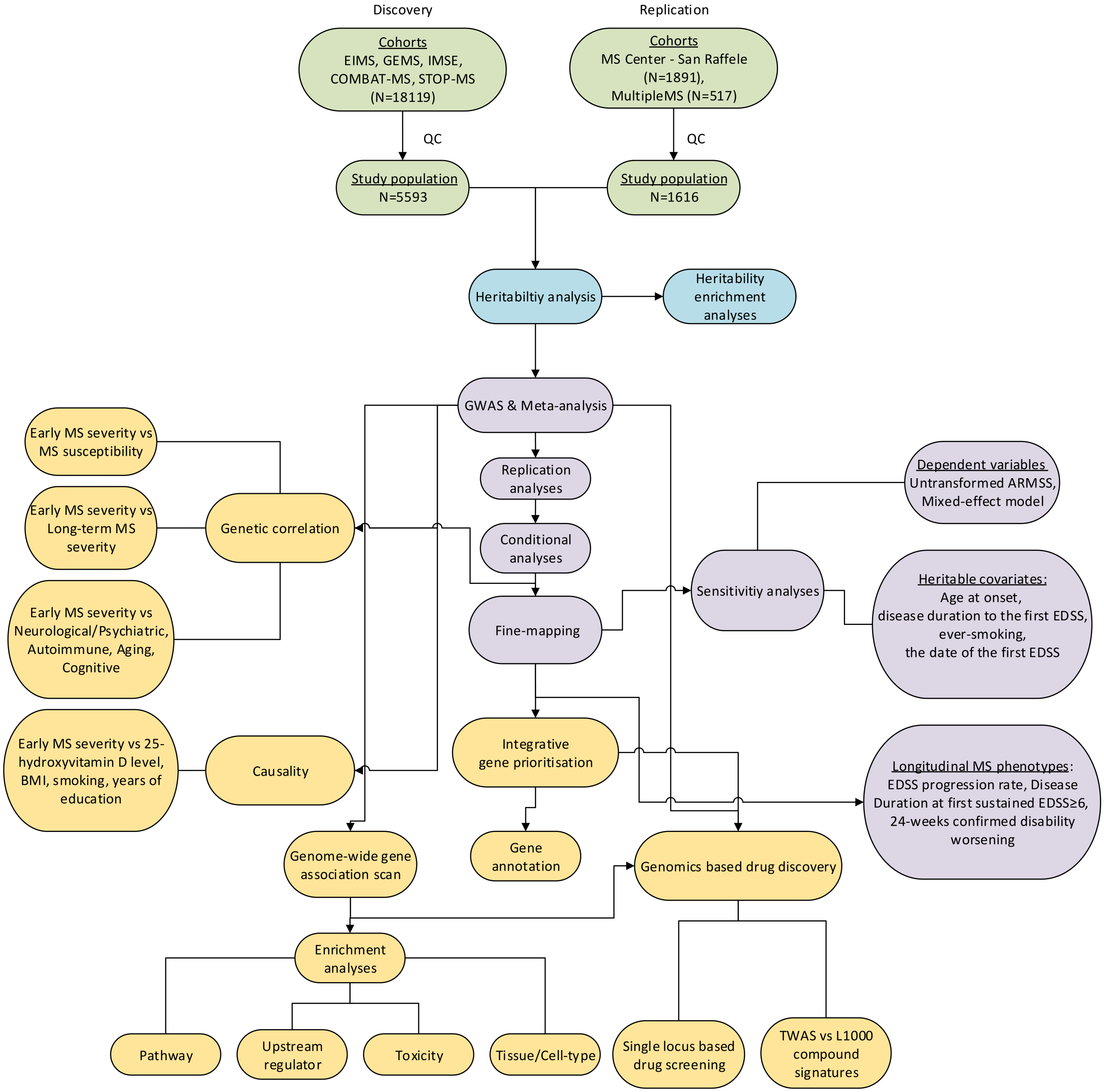


**eFigure 1.** Overview of the Study. Schematic analyses overview displayed by sequence. Green cells indicate Data cleaning and preparation phase, blue denote the pre-GWAS analyses, purple shows the GWAS and its sensitivity analyses, and yellow cells are the downstream analyses performed in this study.

#### eFigure 2.

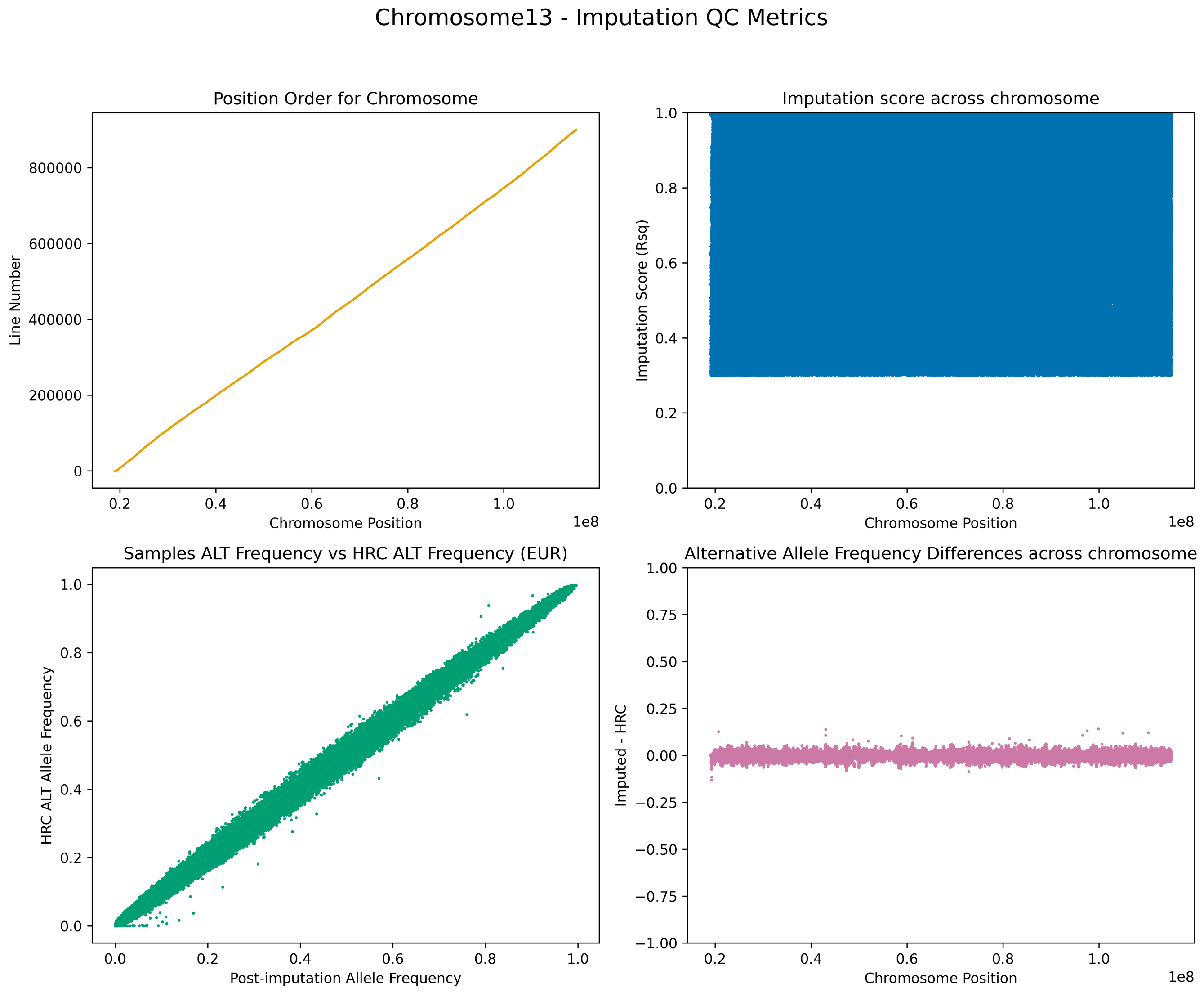


**eFigure 2.** Imputation quality control metrics for chromosome 13 the new Global Screening Array (GSA2) cohort. Four diagnostic panels assess the integrity of genotype imputation: (A) positional continuity across chromosome 13, (B) distribution of imputation quality scores ($R^2$), (C) correlation between sample and European ancestry HRC reference panel alternative allele frequencies, and (D) difference between sample and European ancestry HRC alternative allele frequencies.

#### eFigure 3.

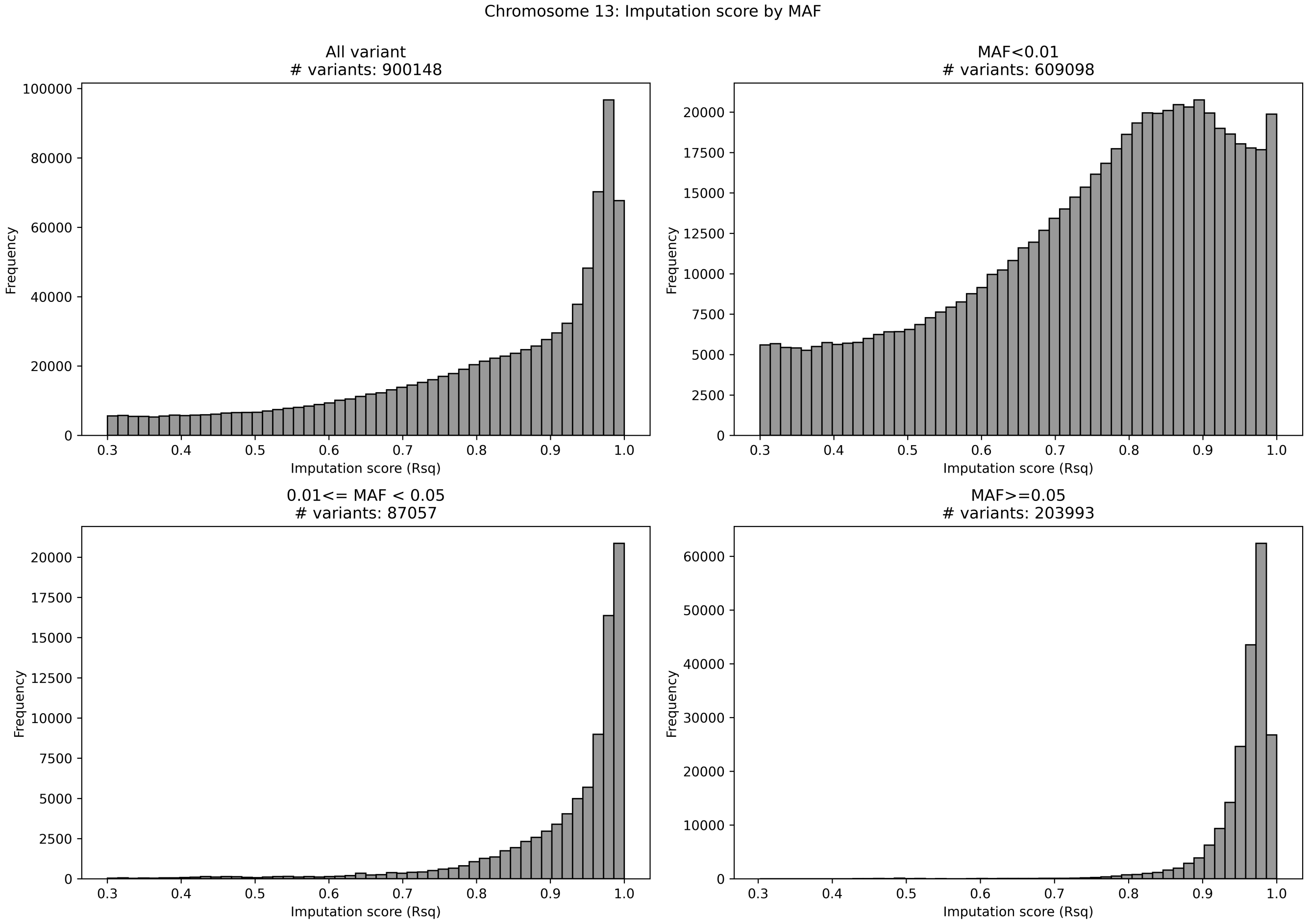


**eFigure 3.** Imputation quality ($R^2$) stratified by minor allele frequency (MAF) bins for chromosome 13 in the GSA2 cohort. Chromosome 13 was presented here as an example for the plots generated for all the chromosomes utilized for this study. (A) All variants; (B) variants with MAF < 0.01; (C) variants with 0.01 <= MAF < 0.05; (D) variants with MAF >= 0.05. # indicates numbers.

#### eFigure 4.

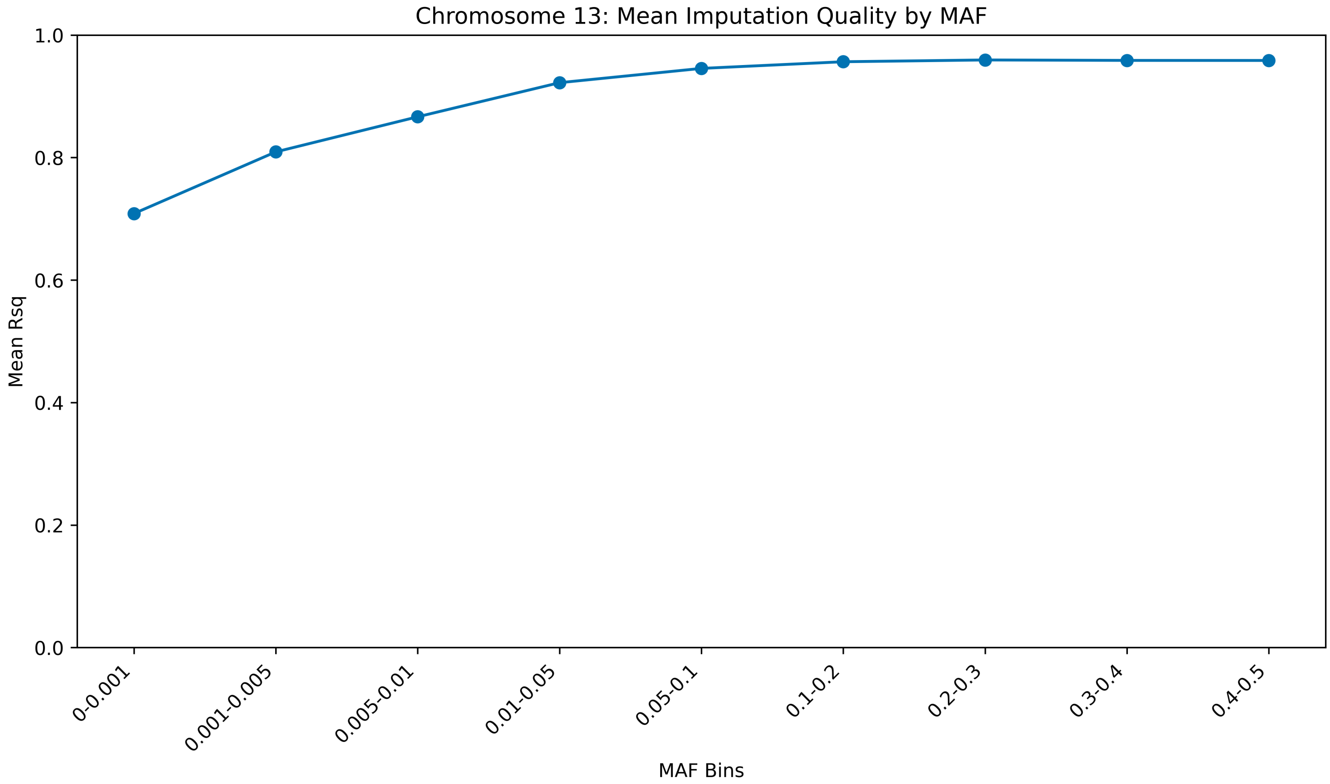


**eFigure 4.** Mean imputation quality ($R^2$) across MAF bins for chromosome 13 in the Swedish multiple sclerosis cohort. Chromosome 13 was presented here as an example for the plots generated for all the chromosomes utilized for this study.

#### eFigure 5.

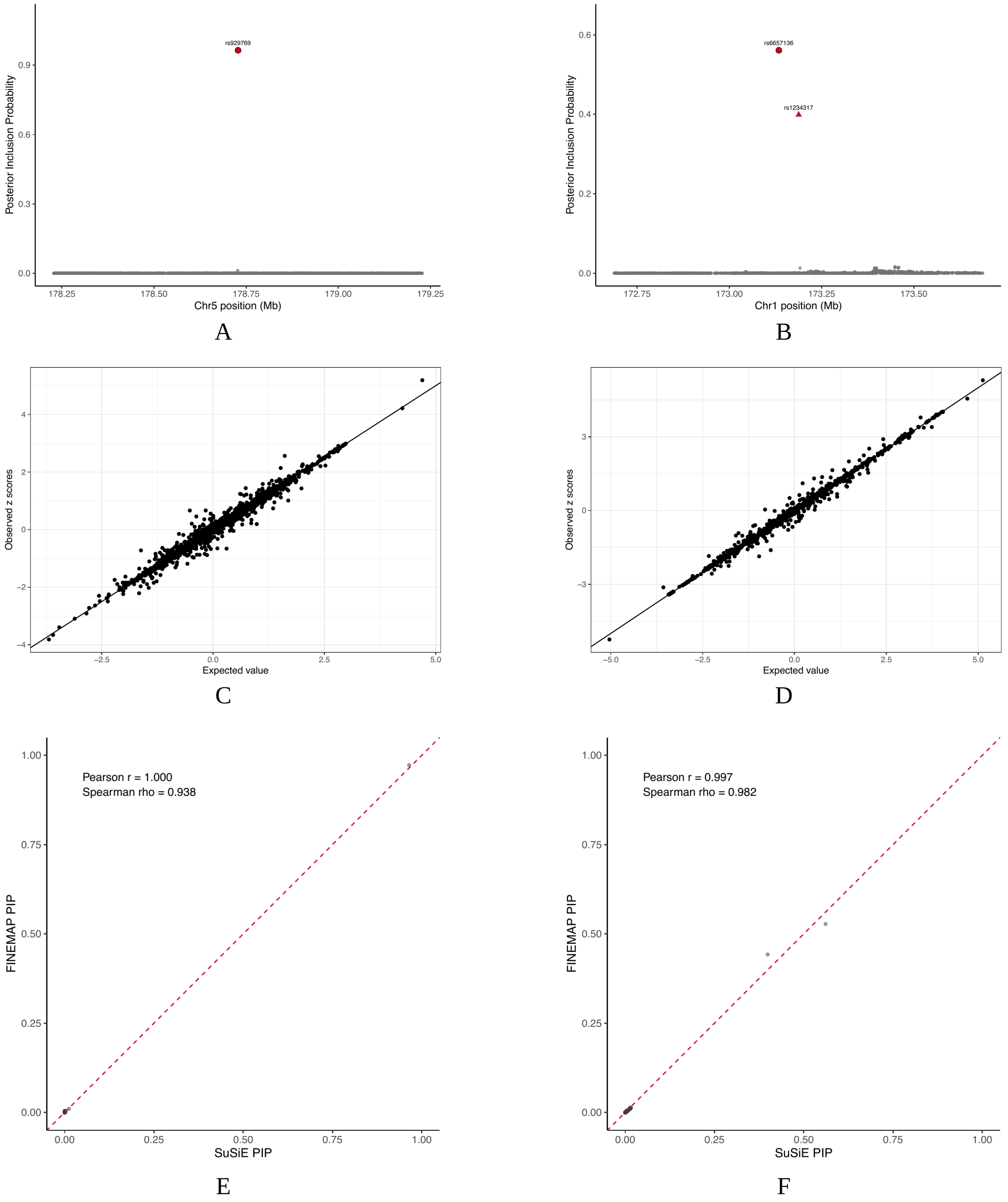


**eFigure 5.** Fine-mapping in replicated loci. (A) Fine-mapping analyses with the posterior inclusion probability (PIP) noted for each replicated locus using SuSiE-RSS in chromosome 5 locus and (B) chromosome 1 locus. (C) The kriging SuSiE-RSS input diagnostic plots checking the relations between the observed GWAS Z scores and the expected Z scores under the sample-size weighted LD matrix for chromosome 5 locus and (D) for chromosome 1 locus. (E) indicates the PIP concordance between SuSiE-RSS and FINEMAP in chromosome 5 locus and (F) for chromosome 1 locus.

**eFigure 6.**

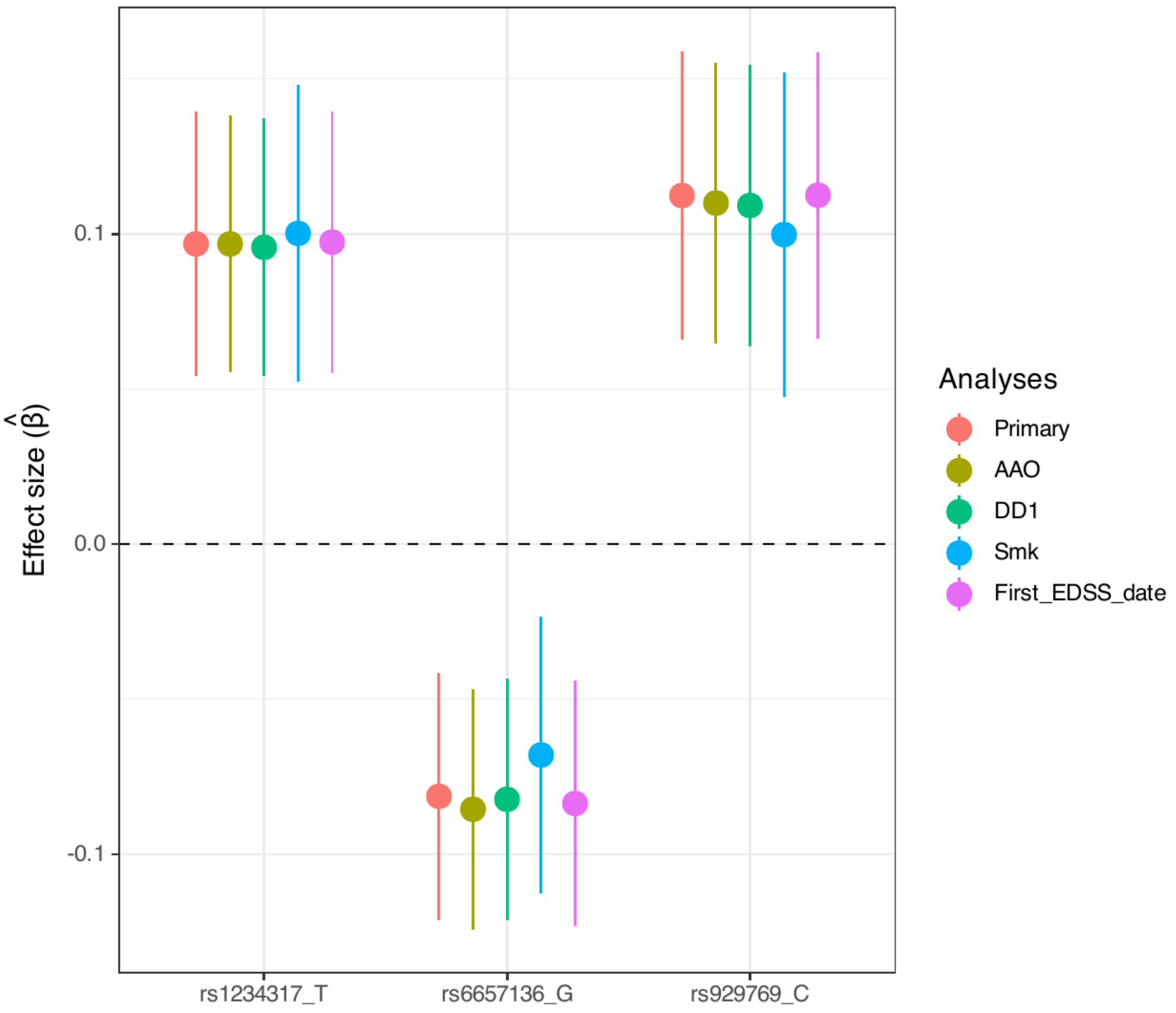


**eFigure 6.** Sensitivity analyses of the early MS severity variants. The effect of each early MS severity allele was adjusted for age at onset (AAO), disease duration from onset to first EDSS measurement (DD1), ever-smoking status prior to the first EDSS measurement (ever vs never smoked; Smk), and date of the first EDSS measurement respectively (First_EDSS_date) and compared to main analysis without any of the mentioned traits adjusted (Primary).

#### eFigure 7.

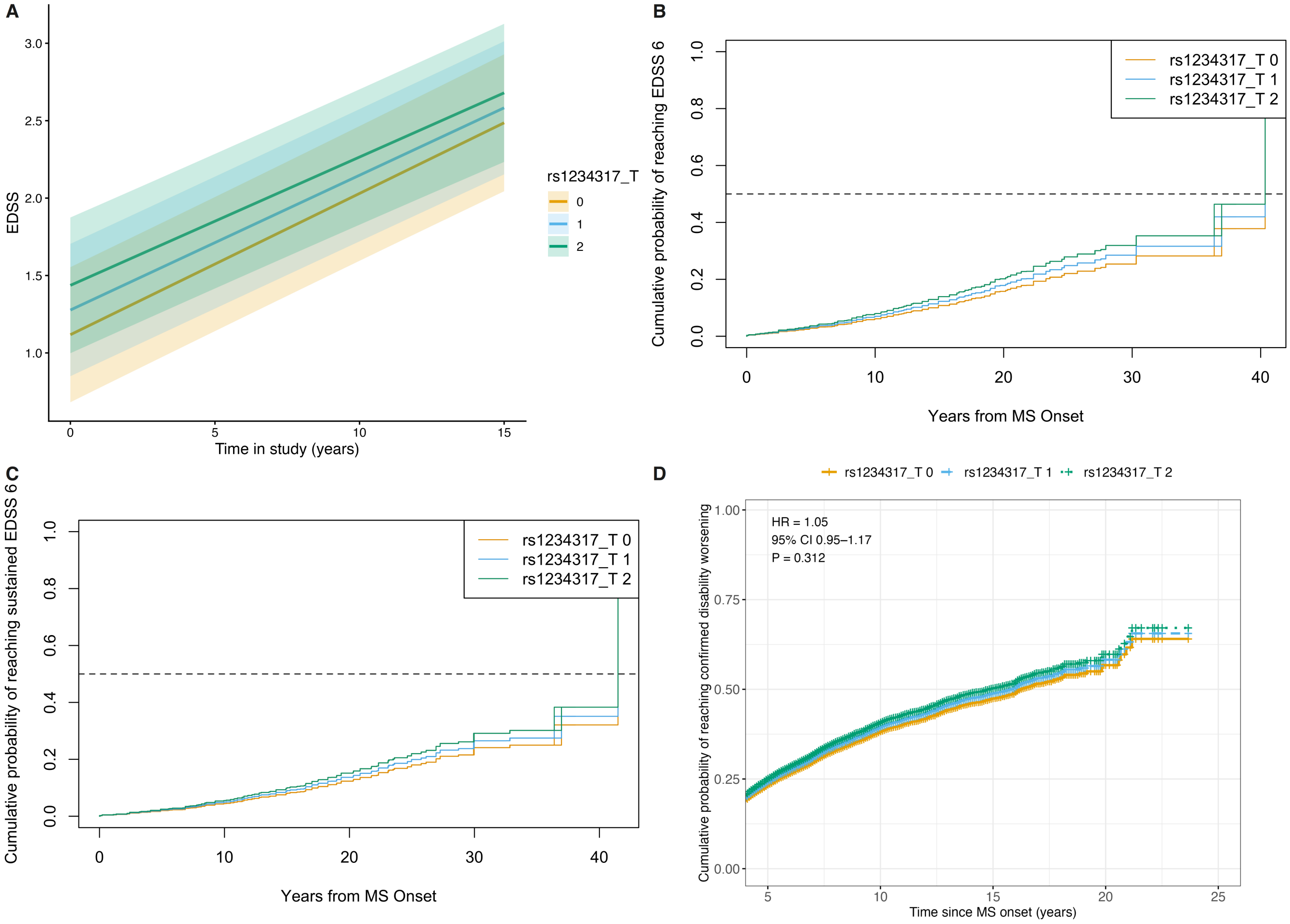


**eFigure 7.** rs1234317 association with time-to-event MS phenotypes. (A) EDSS progression over time ($\beta$ = 0.0980, SE = 0.0429, $P_{geno}$ = 0.0226, $P_{geno\ x\ time}$ = 0.442) (B) Cumulative probability of reaching EDSS 6 over the years from MS onset (HR = 1.15, SE = 0.0504, P = 0.00688). (C) Cumulative probability of reaching sustained EDSS 6 over years from MS onset (HR = 1.12, SE = 0.0532, P = 0.0369). (D) Cumulative probability of experiencing confirmed worsening over the years from MS onset (HR = 1.05, SE = 0.0528, P = 0.312).

#### eFigure 8.

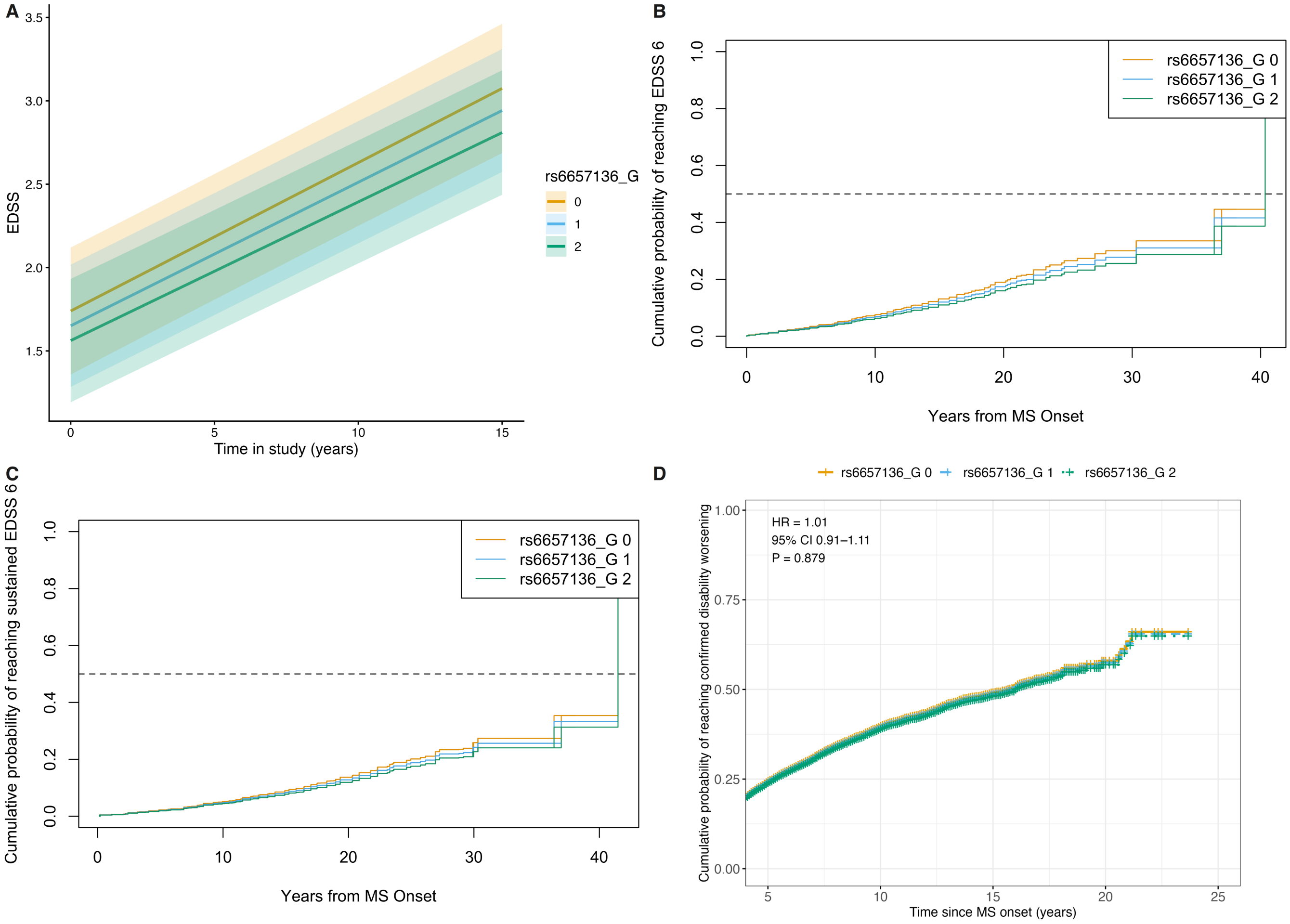


**eFigure 8.** rs6657136 association with time-to-event MS phenotypes. (A) EDSS progression over time ($\beta$ = -0.0887, SE = 0.0405, $P_{geno}$ = 0.0285, $P_{geno\ x\ time}$ = 0.411) (B) Cumulative probability of reaching EDSS 6 over the years from MS onset (HR = 0.91, SE = 0.0452, P = 0.0367). (C) Cumulative probability of reaching sustained EDSS 6 over years from MS onset (HR = 0.927, SE = 0.0519, P = 0.146). (D) Cumulative probability of experiencing confirmed worsening over the years from MS onset (HR = 1.01, SE = 0.0512, P = 0.879).

#### eFigure 9.

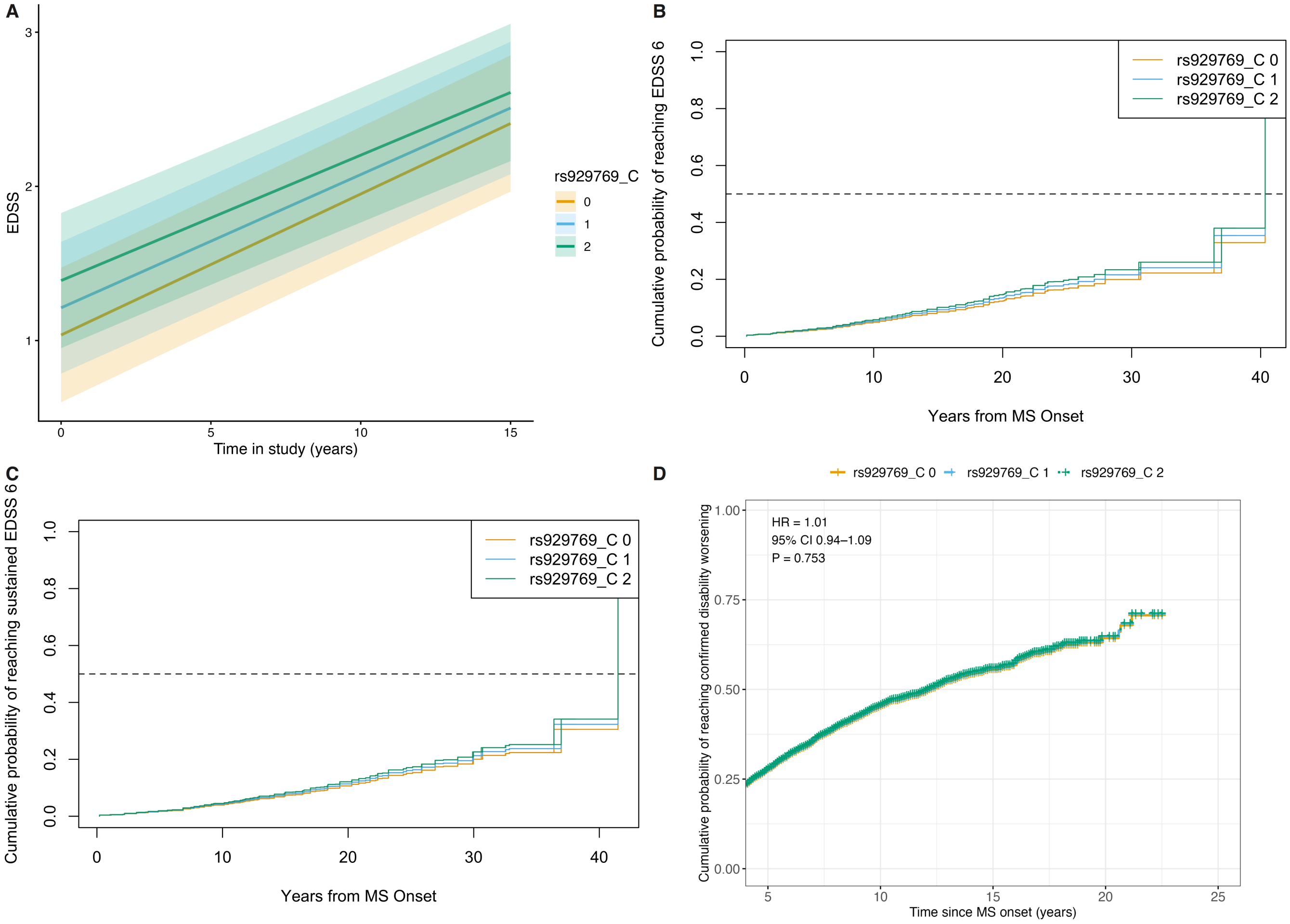


**eFigure 9.** rs929769 association with time-to-event MS phenotypes. (A) EDSS progression over time ($\beta = 0.159$, SE = 0.0472, $P_{geno} = 0.0007$, $P_{geno\ x\ time} = 0.316$). (B) Cumulative probability of reaching EDSS 6 over the years from MS onset (HR = 1.09, SE = 0.0540, P = 0.0947). (C) Cumulative probability of reaching sustained EDSS 6 over years from MS onset (HR = 1.07, SE = 0.0609, P = 0.262). (D) Cumulative probability of experiencing confirmed worsening over the years from MS onset (HR = 1.01, SE = 0.0367, P = 0.753).

#### eFigure 10.

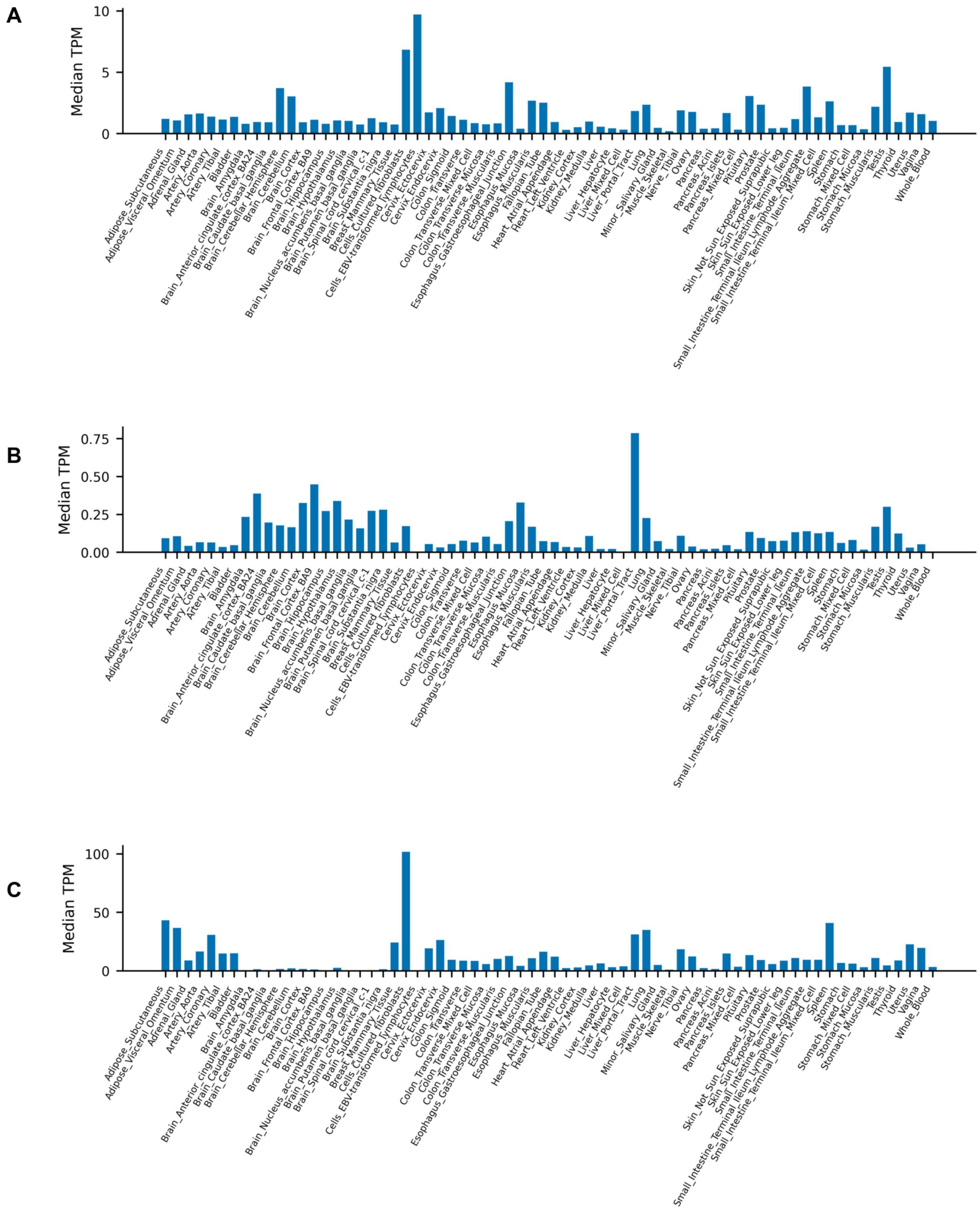


**eFigure 10.** Tissue-specific expressions of the prioritized genes. Tissue-specific expression of (A) TNFSF4, (B) TNFSF18, and (C) ADAMTS2. The expression data is recruited from the GTEx Analysis Release V10 (dbGaP Accession phs000424.v10.p2).

#### eFigure 11.

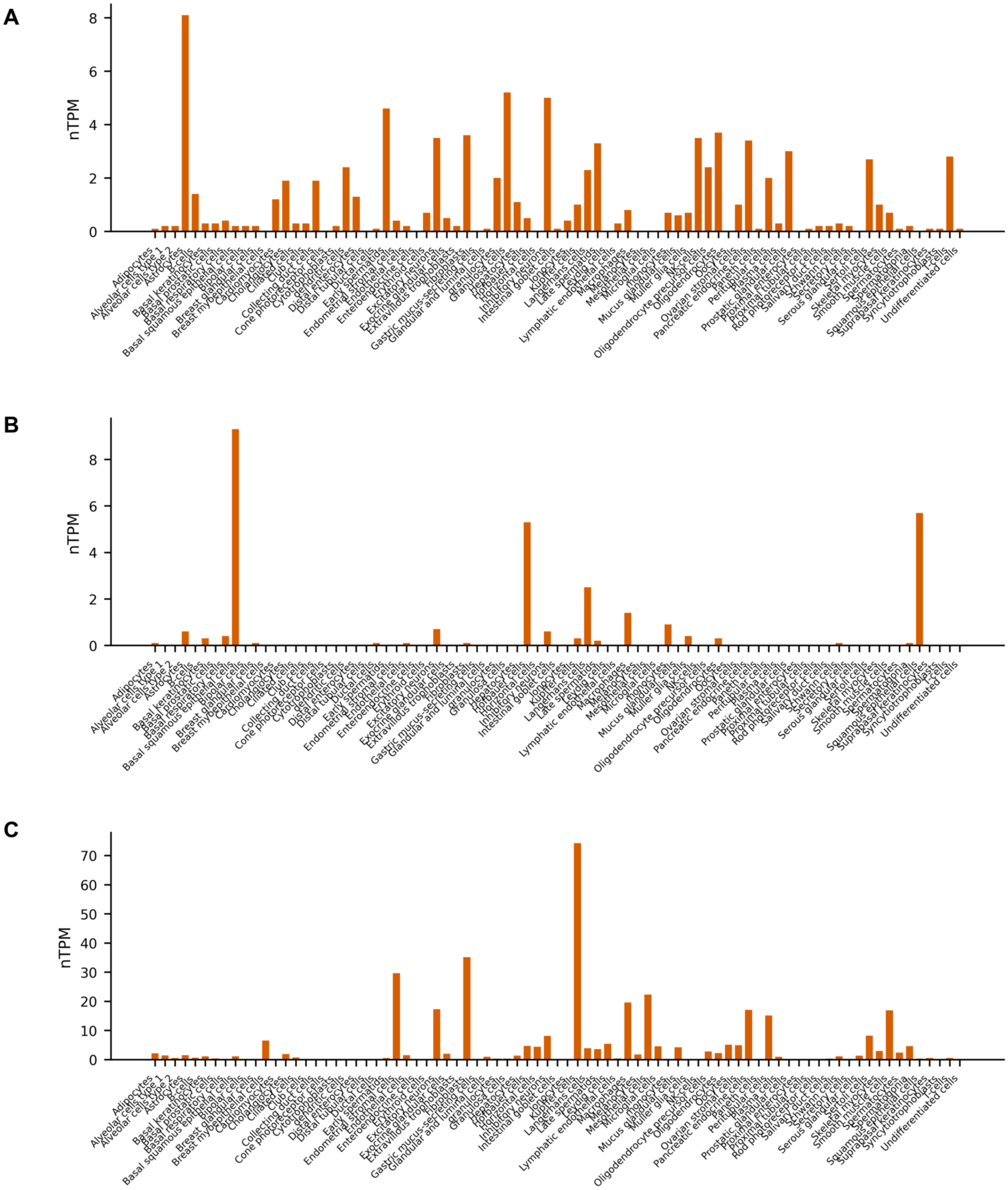


**eFigure 11.** Single cell specific expressions of the prioritized genes. The single cell-type-specific expressions of (A) TNFSF4, (B) TNFSF18, (C) ADAMTS2 using single-cell RNA-sequencing data analyzed from 31 distinct datasets by Human Protein Atlas.

#### eFigure 12.

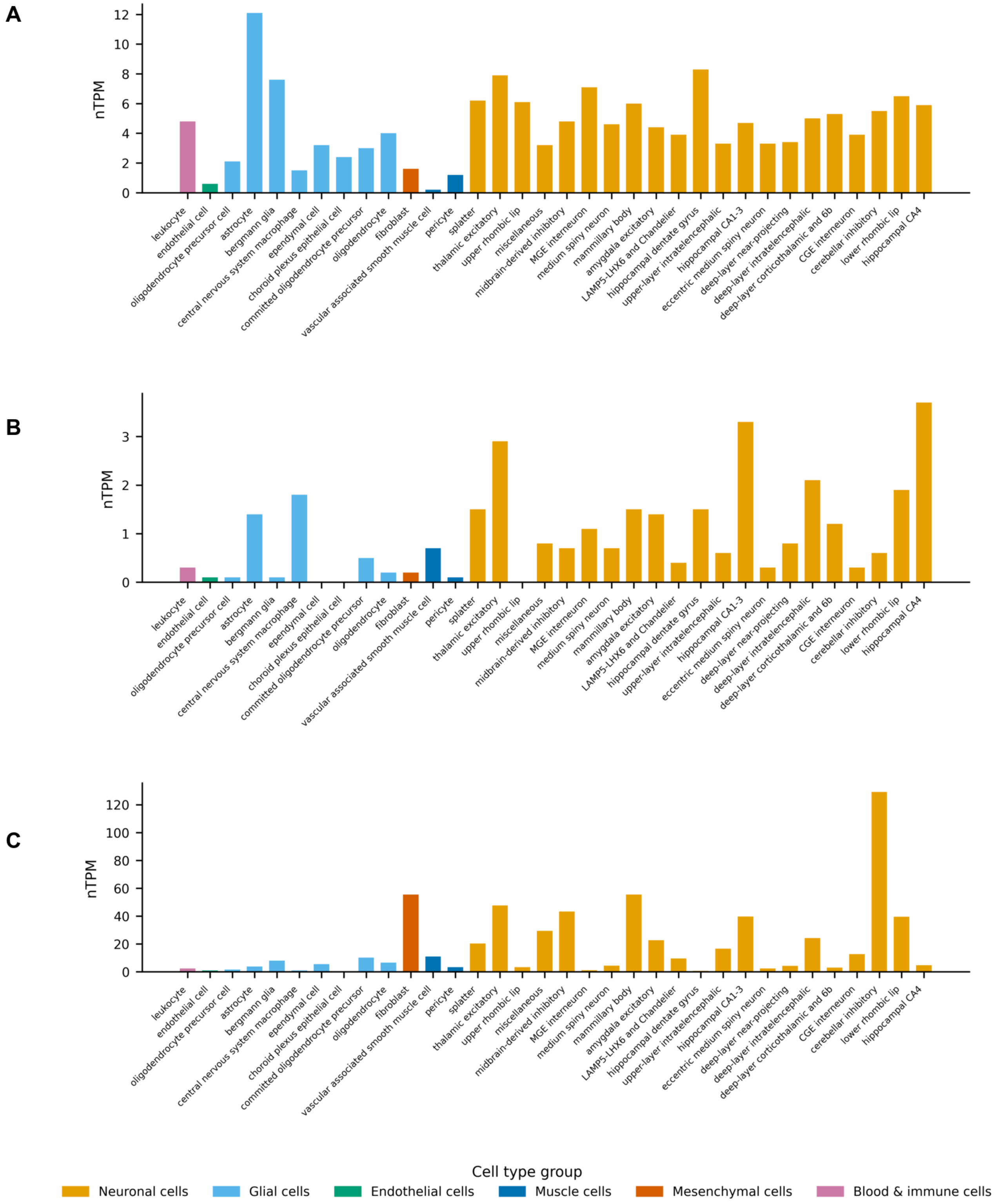


**eFigure 12.** Single-nuclei RNA brain cluster specific expressions of the prioritized genes. The single-nuclei RNA brain cluster specific expressions of (A) TNFSF4, (B) TNFSF18, and (C) ADAMTS2 obtained from the transcript expression level data, summarized per gene, across 34 distinct cell clusters within 11 different brain regions by Human Protein Atlas.

eFigure 13.

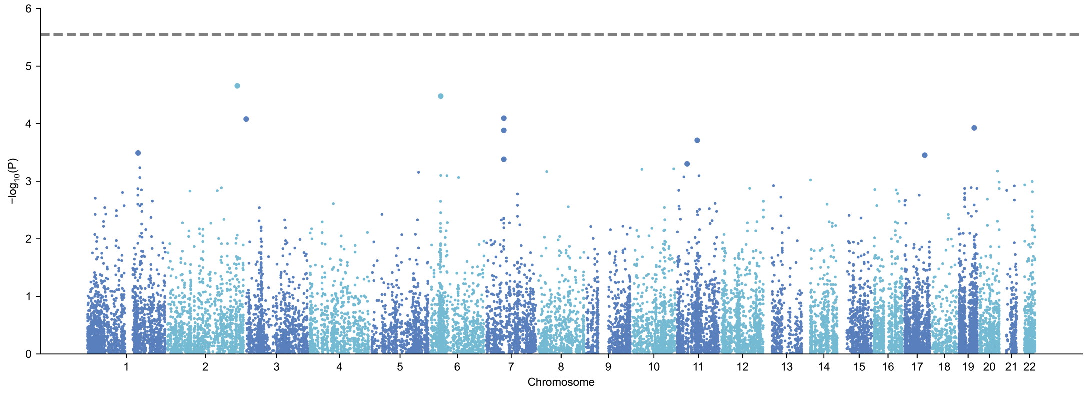


A

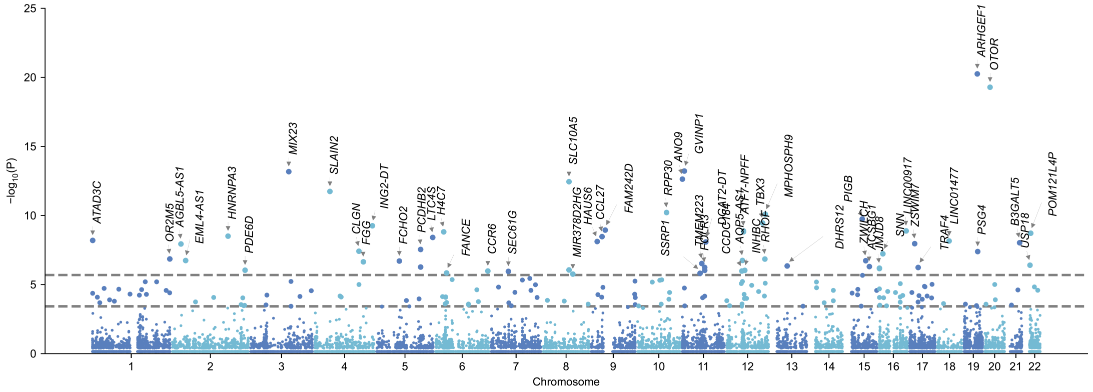


B

**eFigure 13.** Genome-wide gene association scan (GWGAS). Manhattan plots show genome-wide gene association scan conducted using (A) linear model with MAGMA and (B) linear mixed model with LDAK-KVIK-GBAT. Each dot indicates the median position of each gene. The Bonferroni threshold was utilized as genome-wide significance (MAGMA, P = 2.82 x $10^{-6}$; LDAK-KVIK-GBAT, P = 2.03 x $10^{-6}$, and FDR-corrected threshold was used for suggestively significant threshold (LDAK-KVIK-GBAT, P = 3.78 x $10^{-4}$).

#### eFigure 14.

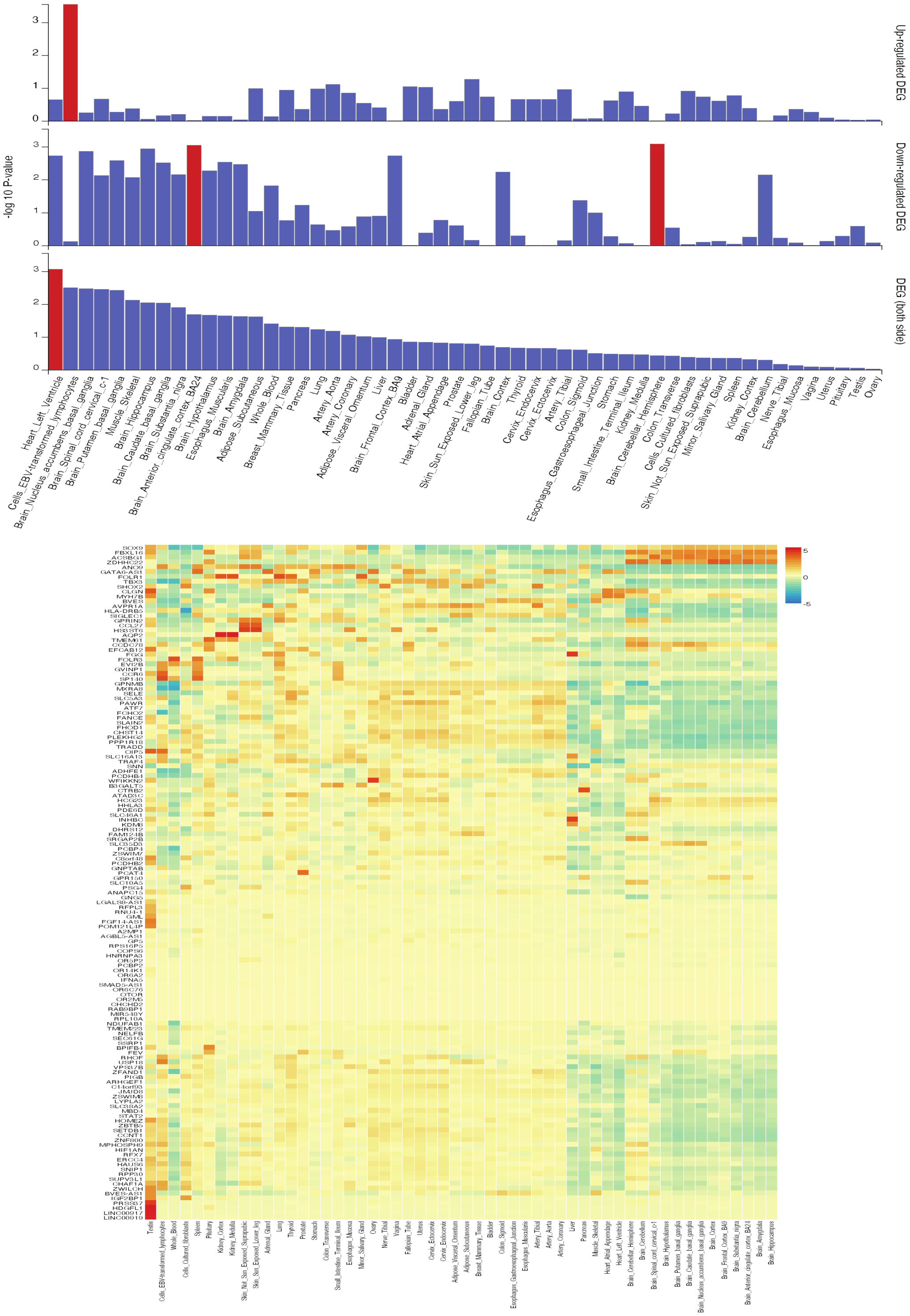


**eFigure 14.** 54 tissue-specific enrichments of pre-computed differentially expressed genes overlapping with early MS severity GWGAS genes. The bar graph demonstrates 54 tissue-specific over-representation analysis between the early MS severity GWGAS genes and pre-computed differentially expressed gene (DEG) sets from GTEx v8. Significant enrichments after Bonferroni correction (P <= 0.05) are indicated by red bars. A normalized expression heatmap of the identical genes across GTEx v8 54 tissues is shown. Genes with higher expression are shown in red and those with lower expression in blue.

#### eFigure 15.

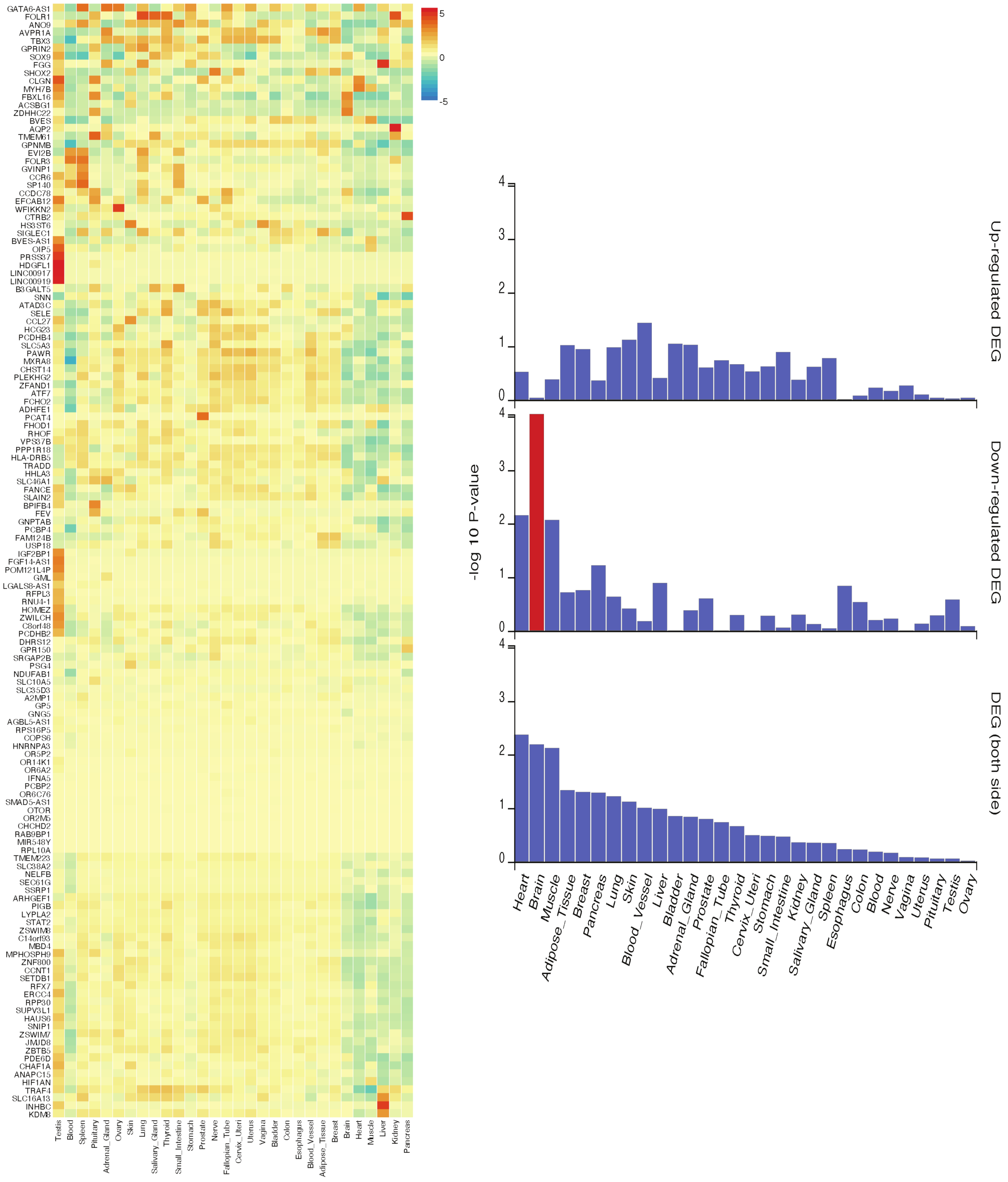


**eFigure 15.** 30 tissue-specific enrichments of pre-computed differentially expressed genes overlapping with early MS severity GWGAS genes. The bar graph demonstrates 30 tissue-specific over-representation analysis between the early MS severity GWGAS genes and pre-computed differentially expressed gene (DEG) sets from GTEx v8. Significant enrichment after Bonferroni correction (P <= 0.05) is indicated by red bars. (B) Normalized expression heatmap of identical genes across GTEx v8 30 general tissues. Genes with higher expression are shown in red and those with lower expression in blue.

### eFigure 16.

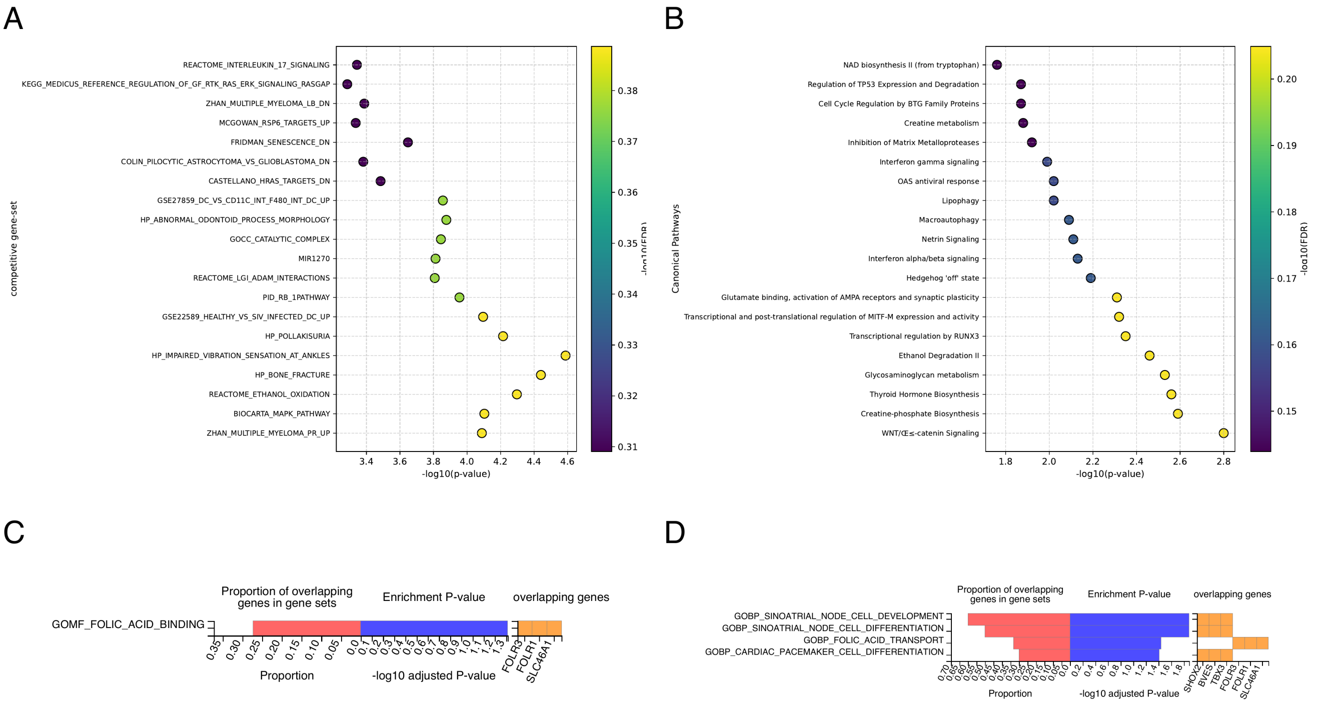


**eFigure 16.** Gene-set analyses. Panels A–D show gene set analyses indicating the top enriched pathways in early MS severity. A, MAGMA competitive gene set analysis. B, IPA canonical pathway analysis. C, Gene Ontology Molecular Function (GOMF) pathway analysis. D, Gene Ontology biological process (GOBP) pathway analysis. C-D pathways are all FDR-significant.

#### eFigure 17.

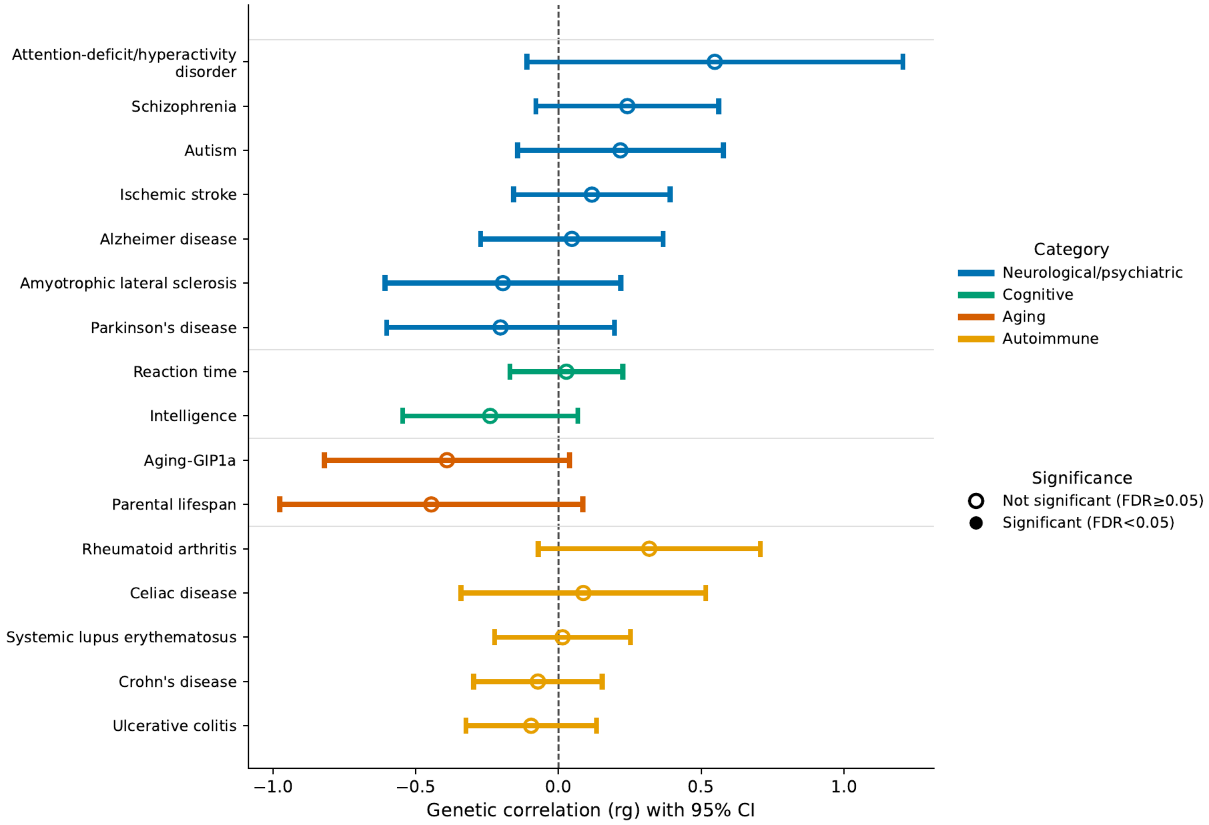


**eFigure 17.** Cross-trait genetic correlations between a range of neurological, psychiatric, autoimmune, aging, and cognitive traits with early MS severity. Neurological/psychiatric traits are colored blue, cognitive traits green, aging red, and autoimmune traits yellow. The insignificant associations have hollow circles.

#### eFigure 18.

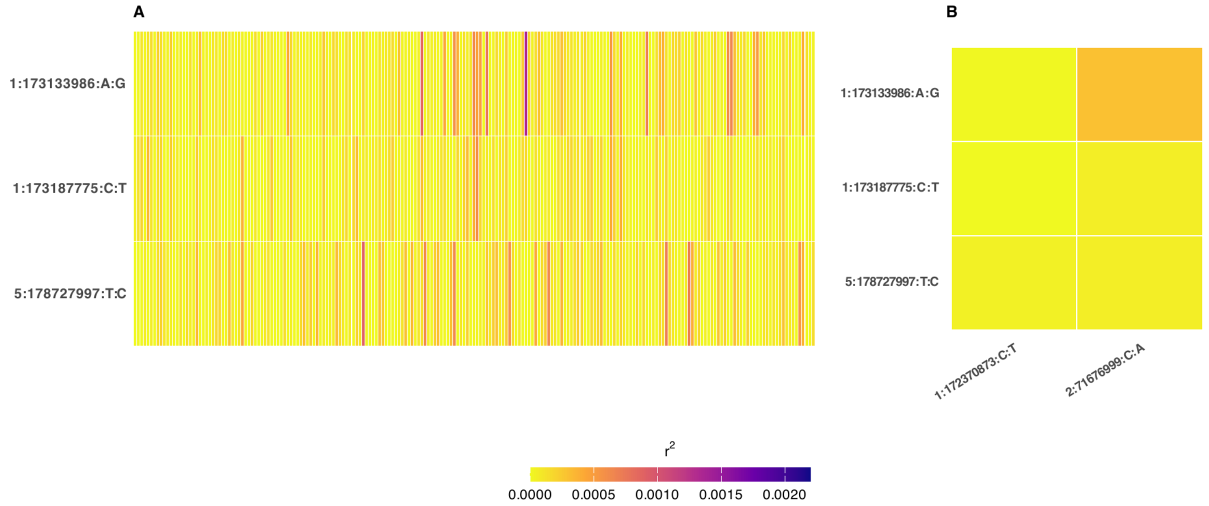


**eFigure 18.** Linkage disequilibrium ($r^2$) between the early MS severity and late MS severity and MS susceptibility SNPs. The early MS severity SNPs are noted on y-axes. (A) The linkage disequilibrium (LD) blocks between the early MS severity and MS susceptibility SNPs. (B) shows the LD relationships the early severity SNPs and the late MS severity SNPs.

#### eFigure 19.

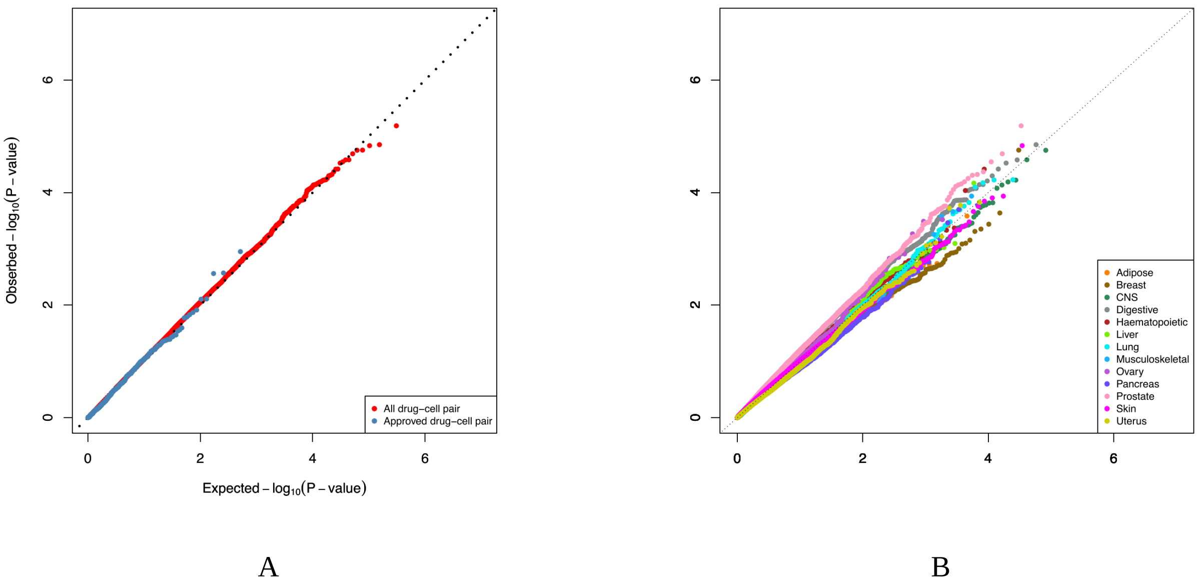


**eFigure 19.** The QQ plots for Drug-cell inverse expression signatures screening. (A) and (B) are QQ plots of -$\log_{10}$(P-value) indicating the negative spearman correlations between the predicted genetically regulated expression and compound-regulated gene expression profiles. The diagonal line indicates a null correlation. The x-axes denote the expected -$\log_{10}$(P-value), and the y-axes indicate the observed -$\log_{10}$(P-value). (A) includes all drug compounds and approved drug compound correlations. (B) has tissue-specific correlations.

#### eReferences


1. Hedström AK, Bäärnhielm M, Olsson T, Alfredsson L. Tobacco smoking, but not Swedish snuff use, increases the risk of multiple sclerosis. *Neurology*. 2009;73(9):696-701. doi:10.1212/wnl.0b013e3181b59c40

2. Hedström AK, Hillert J, Olsson T, Alfredsson L. Nicotine might have a protective effect in the etiology of multiple sclerosis. *Multiple Sclerosis Journal*. 2013;19(8):1009-1013. doi:10.1177/1352458512471879

3. Holmén C, Piehl F, Hillert J, et al. The "Immunomodulation and Multiple Sclerosis Epidemiology" (IMSE) study : a Swedish nationwide pharmaco-epidemiological and genetic study focused on long-term safety and efficacy of natalizumab (Tysabri). *European Journal of Neurology*. 2012;19:751-751.

4. F P, P A, S V, et al. COMBAT-MS: A Population-Based Observational Cohort Study Addressing the Benefit-Risk Balance of Multiple Sclerosis Therapies Compared with Rituximab. *PubMed*. Accessed February 4, 2026. https://pubmed.ncbi.nlm.nih.gov/38923558/

5. Khademi M, Dring A, Gilthorpe JD, et al. Intense Inflammation and Nerve Damage in Early Multiple Sclerosis Subsides at Older Age: A Reflection by Cerebrospinal Fluid Biomarkers. *PLoS ONE*. 2013;8(5). doi:10.1371/journal.pone.0063172

6. Hillert J, Stawiarz L. The Swedish MS registry – clinical support tool and scientific resource. *Acta Neurol Scand*. 2015;132(S199):11-19. doi:10.1111/ane.12425

7. Uibel P, Trogu F, Flaskamp M, et al. Therapy Choices and Outcomes in Newly Diagnosed Multiple Sclerosis: The PanEuropean MultipleMS Study. *Neurology: Open Access*. Published online In press.

8. Thompson AJ, Banwell BL, Barkhof F, et al. Diagnosis of multiple sclerosis: 2017 revisions of the McDonald criteria. Accessed May 28, 2026. https://www.thelancet.com/journals/laneur/article/PIIS1474-4422(17)30470-2/abstract

9. Kurtzke JF. Rating neurologic impairment in multiple sclerosis. *Neurology*. 1983;33(11):1444-1444. doi:10.1212/wnl.33.11.1444

10. Ramo-Tello C, Blanco Y, Brieva L, et al. Recommendations for the Diagnosis and Treatment of Multiple Sclerosis Relapses. *Journal of Personalized Medicine*. 2021;12(1):6-6. doi:10.3390/jpm12010006

11. Manouchehrinia A, Westerlind H, Kingwell E, et al. Age Related Multiple Sclerosis Severity Score: Disability ranked by age. *Mult Scler*. 2017;23(14):1938-1946. doi:10.1177/1352458517690618

12. International Multiple Sclerosis Genetics Consortium, Harroud A, Stridh P, et al. Locus for severity implicates CNS resilience in progression of multiple sclerosis. *Nature*. 2023;619(7969):323-331. doi:10.1038/s41586-023-06250-x

13. Sheng X, Xia L, Cahoon JL, et al. Inverted genomic regions between reference genome builds in humans impact imputation accuracy and decrease the power of association testing. Accessed October 22, 2025. https://www.cell.com/hgg-advances/abstract/S2666-2477(22)00076-8

14. Chang C, Chow CC, Tellier LC, Vattikuti S, Purcell S, Lee JJ. Second-generation PLINK: rising to the challenge of larger and richer datasets. *GigaScience*. 2015;4(1). doi:10.1186/s13742-015-0047-8

15. Privé F, Luu K, Blum MGB, McGrath JJ, Vilhjálmsson BJ. Efficient toolkit implementing best practices for principal component analysis of population genetic data. *Bioinformatics*. 2020;36(16):4449-4457. doi:10.1093/bioinformatics/btaa520

16. Loh PR, Danecek P, Palamara PF, et al. Reference-based phasing using the Haplotype Reference Consortium panel. *Nature Genetics*. 2016;48(11):1443-1448. doi:10.1038/ng.3679

17. McCarthy S, Das S, Kretzschmar WW, et al. A reference panel of 64,976 haplotypes for genotype imputation. *Nature Genetics*. 2016;48(10):1279-1283. doi:10.1038/ng.3643

18. Das S, Forer L, Schönherr S, et al. Next-generation genotype imputation service and methods. *Nature Genetics*. 2016;48(10):1284-1287. doi:10.1038/ng.3656

19. The International Multiple Sclerosis Genetics Consortium & The Wellcome Trust Case Control Consortium 2. Genetic risk and a primary role for cell-mediated immune mechanisms in multiple sclerosis. *Nature*. 2011;476(7359):214-219. doi:10.1038/nature10251

20. Purcell S, Neale BM, Todd-Brown KE, et al. PLINK: A Tool Set for Whole-Genome Association and Population-Based Linkage Analyses. *The American Journal of Human Genetics*. 2007;81(3):559-575. doi:10.1086/519795

21. Zou Y, Carbonetto P, Wang G, Stephens M. Fine-mapping from summary data with the “Sum of Single Effects” model. doi:10.1371/journal.pgen.1010299

22. Benner C, Spencer CCA, Havulinna AS, Salomaa V, Ripatti S, Pirinen M. FINEMAP: efficient variable selection using summary data from genome-wide association studies. *Bioinformatics*. 2016;32(10):1493-1501. doi:10.1093/bioinformatics/btw018

23. Kuhlmann T, Moccia M, Coetzee T, et al. Multiple sclerosis progression: time for a new mechanism-driven framework. *The Lancet Neurology*. 2023;22(1):78-88. doi:10.1016/S1474-4422(22)00289-7

24. 1000 Genomes Project Consortium, Auton A, Brooks LD, et al. A global reference for human genetic variation. *Nature*. 2015;526(7571):68-74. doi:10.1038/nature15393

25. Harrison PW, Amode MR, Austine-Orimoloye O, et al. Ensembl 2024. *Nucleic Acids Research*. 2023;52(D1):D891-D899. doi:10.1093/nar/gkad1049

26. Hinrichs AS. The UCSC Genome Browser Database: update 2006. *Nucleic Acids Research*. 2006;34(90001):D590-D598. doi:10.1093/nar/gkj144

27. Wang K, Li M, Hakonarson H. ANNOVAR: functional annotation of genetic variants from high-throughput sequencing data. *Nucleic Acids Res*. 2010;38(16):e164. doi:10.1093/nar/gkq603

28. Aguet F, Anand S, Ardlie K, et al. The GTEx Consortium atlas of genetic regulatory effects across human tissues. *Science*. 2020;369(6509):1318-1330. doi:10.1126/science.aaz1776

29. Kerimov N, Hayhurst J, Peikova K, et al. A compendium of uniformly processed human gene expression and splicing quantitative trait loci. *Nature Genetics*. 2021;53(9):1290-1299. doi:10.1038/s41588-021-00924-w

30. Kerimov N, Tambets R, Hayhurst JD, et al. eQTL Catalogue 2023: New datasets, X chromosome QTLs, and improved detection and visualisation of transcript-level QTLs. doi:10.1371/journal.pgen.1010932

31. Schmitt AD, Hu M, Jung I, et al. A compendium of chromatin contact maps reveals spatially active regions in the human genome. *Cell Rep*. 2016;17(8):2042-2059. doi:10.1016/j.celrep.2016.10.061

32. Nasser J, Bergman DT, Fulco CP, et al. Genome-wide enhancer maps link risk variants to disease genes. *Nature*. 2021;593(7858):238-243. doi:10.1038/s41586-021-03446-x

33. Javierre BM, Burren OS, Wilder SP, et al. Lineage-specific genome architecture links enhancers and non-coding disease variants to target gene promoters. *Cell*. 2016;167(5):1369-1384.e19. doi:10.1016/j.cell.2016.09.037

34. Mountjoy E, Schmidt EM, Carmona M, et al. An open approach to systematically prioritize causal variants and genes at all published human GWAS trait-associated loci. *Nature Genetics*. 2021;53(11):1527-1533. doi:10.1038/s41588-021-00945-5

35. Ghoussaini M, Mountjoy E, Carmona M, et al. Open Targets Genetics: systematic identification of trait-associated genes using large-scale genetics and functional genomics. *Nucleic Acids Res*. 2020;49(D1):D1311-D1320. doi:10.1093/nar/gkaa840

36. Ochoa D, Hercules A, Carmona M, et al. Open Targets Platform: supporting systematic drug–target identification and prioritisation. *Nucleic Acids Res*. 2021;49(D1):D1302-D1310. doi:10.1093/nar/gkaa1027

37. West CE, Karim M, Falaguera MJ, et al. Integrative GWAS and co-localisation analysis suggests novel genes associated with age-related multimorbidity. *Sci Data*. 2023;10(1). doi:10.1038/s41597-023-02513-4

38. Gazal S, Weissbrod O, Hormozdiari F, et al. Combining SNP-to-gene linking strategies to identify disease genes and assess disease omnigenicity. *Nature Genetics*. 2022;54(6):827-836. doi:10.1038/s41588-022-01087-y

39. Ma S, Wang C, Khan A, et al. BIGKnock: fine-mapping gene-based associations via knockoff analysis of biobank-scale data. *Genome Biol*. 2023;24(1). doi:10.1186/s13059-023-02864-6

40. Fromer M, Roussos P, Sieberts SK, et al. Gene expression elucidates functional impact of polygenic risk for schizophrenia. *Nature Neuroscience*. 2016;19(11):1442-1453. doi:10.1038/nn.4399

41. Gusev A, Ko A, Shi H, et al. Integrative approaches for large-scale transcriptome-wide association studies. *Nature Genetics*. 2016;48(3):245-252. doi:10.1038/ng.3506

42. Giambartolomei C, Vukcevic D, Schadt EE, et al. Bayesian Test for Colocalisation between Pairs of Genetic Association Studies Using Summary Statistics. Williams SM, ed. *PLoS Genet*. 2014;10(5):e1004383. doi:10.1371/journal.pgen.1004383

43. Wallace C. A more accurate method for colocalisation analysis allowing for multiple causal variants. Cordell HJ, ed. *PLoS Genet*. 2021;17(9):e1009440. doi:10.1371/journal.pgen.1009440

44. Wainberg M, Sinnott-Armstrong N, Mancuso N, et al. Opportunities and challenges for transcriptome-wide association studies. *Nat Genet*. 2019;51(4):592-599. doi:10.1038/s41588-019-0385-z

45. Zhao S, Crouse W, Qian S, Luo K, Stephens M, He X. Adjusting for genetic confounders in transcriptome-wide association studies improves discovery of risk genes of complex traits. *Nat Genet*. 2024;56(2):336-347. doi:10.1038/s41588-023-01648-9

46. Siletti K, Hodge R, Mossi Albiach A, et al. Transcriptomic diversity of cell types across the adult human brain. *Science*. 2023;382(6667):eadd7046. doi:10.1126/science.add7046

47. Digre A, Lindskog C. The human protein atlas—Integrated omics for single cell mapping of the human proteome. *Protein Science*. 2023;32(2). doi:10.1002/pro.4562

48. De Leeuw CA, Mooij JM, Heskes T, Posthuma D. MAGMA: Generalized Gene-Set Analysis of GWAS Data. Tang H, ed. *PLoS Comput Biol*. 2015;11(4):e1004219. doi:10.1371/journal.pcbi.1004219

49. Hof JP, Speed D. LDAK-KVIK performs fast and powerful mixed-model association analysis of quantitative and binary phenotypes. *Nature Genetics*. 2025;57(9):2116-2123. doi:10.1038/s41588-025-02286-z

50. Berrandou TE, Balding D, Speed D. LDAK-GBAT: Fast and powerful gene-based association testing using summary statistics. *The American Journal of Human Genetics*. 2023;110(1):23-29. doi:10.1016/j.ajhg.2022.11.010

51. Tiane A, Schepers M, Reijnders RA, et al. From methylation to myelination: epigenomic and transcriptomic profiling of chronic inactive demyelinated multiple sclerosis lesions. *Acta Neuropathol*. 2023;146(2):283-299. doi:10.1007/s00401-023-02596-8

52. Humphries ESA, Dart C. Neuronal and Cardiovascular Potassium Channels as Therapeutic Drug Targets. *J Biomol Screen*. 2015;20(9):1055-1073. doi:10.1177/1087057115601677

53. Skavinska O, Rossokha Z, Fishchuk L, Gorovenko N. *RFC* and *VDR* -mediated genetic regulation of brain folate transport in patients with multiple sclerosis. *Human Gene*. 2025;44:201399. doi:10.1016/j.humgen.2025.201399

54. Aşcı AE, Orhan G, Karahalil B. Genetic variants of folate metabolism and the risk of multiple sclerosis. *Neurological Research*. 2024;46(6):544-552. doi:10.1080/01616412.2024.2337519

55. Masanneck L, Eichler S, Vogelsang A, et al. The STING-IFN-β-Dependent Axis Is Markedly Low in Patients with Relapsing-Remitting Multiple Sclerosis. *Int J Mol Sci*. 2020;21(23):9249. doi:10.3390/ijms21239249

56. Ke Y, Wang C, Zhang J, et al. The Role of PARPs in Inflammation—And Metabolic—Related Diseases: Molecular Mechanisms and Beyond. *Cells*. 2019;8(9):1047. doi:10.3390/cells8091047

57. Woo MS, Mayer C, Binkle-Ladisch L, et al. STING orchestrates the neuronal inflammatory stress response in multiple sclerosis. *Cell*. 2024;187(15):4043-4060.e30. doi:10.1016/j.cell.2024.05.031

58. Haghgouie T, Shirafkan M, Shomali N. The Impact of Insulin-Like Growth Factor on the Regeneration of Myelin Sheath. *Biomedical Research Bulletin*. 2024;2(4):204-208. doi:10.34172/biomedrb.2024.28

59. International Multiple Sclerosis Genetics Consortium, Patsopoulos NA, Baranzini SE, et al. Multiple sclerosis genomic map implicates peripheral immune cells and microglia in susceptibility. *Science*. 2019;365(6460):eaav7188. doi:10.1126/science.aav7188

60. Choi SW, Mak TSH, O'Reilly PF. Tutorial: a guide to performing polygenic risk score analyses. *Nat Protoc*. 2020;15(9):2759-2772. doi:10.1038/s41596-020-0353-1

61. Cerezo M, Sollis E, Ji Y, et al. The NHGRI-EBI GWAS Catalog: standards for reusability, sustainability and diversity. *Nucleic Acids Res*. 2025;53(D1):D998-D1005. doi:10.1093/nar/gkae1070

62. Bycroft C, Freeman C, Petkova D, et al. The UK Biobank resource with deep phenotyping and genomic data. *Nature*. 2018;562(7726):203-209. doi:10.1038/s41586-018-0579-z

63. Kurki MI, Karjalainen J, Palta P, et al. FinnGen provides genetic insights from a well-phenotyped isolated population. *Nature*. 2023;613(7944):508-518. doi:10.1038/s41586-022-05473-8

64. Jacobs BM, Noyce AJ, Giovannoni G, Dobson R. BMI and low vitamin D are causal factors for multiple sclerosis. *Neurology Neuroimmunology & Neuroinflammation*. 2020;7(2). doi:10.1212/nxi.0000000000000662

65. Jareebi M, Shami MO, Gharawi NF, et al. Assessing the causal relationship between modifiable risk factors, including vitamin D, and the risk of Multiple Sclerosis (MS) using two-sample Mendelian randomization analysis. *International Journal of Medicine in Developing Countries*. Published online January 2023:1-1. doi:10.24911/ijmdc.51-1684898756

66. Bjørnevik K, Riise T, Cortese M, et al. Level of education and multiple sclerosis risk after adjustment for known risk factors: The EnvIMS study. *Multiple Sclerosis Journal*. 2015;22(1):104-111. doi:10.1177/1352458515579444

67. Verbanck M, Chen CY, Neale BM, Do R. Detection of widespread horizontal pleiotropy in causal relationships inferred from Mendelian randomization between complex traits and diseases. *Nature Genetics*. 2018;50(5):693-698. doi:10.1038/s41588-018-0099-7

68. Hemani G, Bowden J, Smith GD. Evaluating the potential role of pleiotropy in Mendelian randomization studies. *Human Molecular Genetics*. 2018;27. doi:10.1093/hmg/ddy163

69. Morris JH, Soman K, Akbas RE, et al. The scalable precision medicine open knowledge engine (SPOKE): a massive knowledge graph of biomedical information. *Bioinformatics*. 2023;39(2):btad080. doi:10.1093/bioinformatics/btad080

70. Zdrazil B, Felix E, Hunter F, et al. The ChEMBL Database in 2023: a drug discovery platform spanning multiple bioactivity data types and time periods. *Nucleic Acids Res*. 2024;52(D1):D1180-D1192. doi:10.1093/nar/gkad1004

71. Liu T, Hwang L, Burley SK, et al. BindingDB in 2024: a FAIR knowledgebase of protein-small molecule binding data. *Nucleic Acids Res*. 2025;53(D1):D1633-D1644. doi:10.1093/nar/gkae1075

72. Liu X, Zhu F, Ma X, et al. The Therapeutic Target Database: an Internet resource for the primary targets of approved, clinical trial and experimental drugs. *Expert Opinion on Therapeutic Targets*. 2011;15(8):903-912. doi:10.1517/14728222.2011.586635

73. Whirl-Carrillo M, Huddart R, Gong L, et al. An Evidence-Based Framework for Evaluating Pharmacogenomics Knowledge for Personalized Medicine. *Clinical Pharmacology & Therapeutics*. 2021;110(3):563-572. doi:10.1002/cpt.2350

74. Knox C, Wilson M, Klinger CM, et al. DrugBank 6.0: the DrugBank Knowledgebase for 2024. *Nucleic Acids Res*. 2024;52(D1):D1265-D1275. doi:10.1093/nar/gkad976

75. Reay WR, Cairns MJ. Advancing the use of genome-wide association studies for drug repurposing. *Nature Reviews Genetics*. 2021;22(10):658-671. doi:10.1038/s41576-021-00387-z

76. Mancuso N, Freund MK, Johnson R, et al. Probabilistic fine-mapping of transcriptome-wide association studies. *Nature Genetics*. 2019;51(4):675-682. doi:10.1038/s41588-019-0367-1

77. Namba S, Konuma T, Wu KH, Zhou W, Okada Y. A practical guideline of genomics-driven drug discovery in the era of global biobank meta-analysis. Accessed September 8, 2025. https://www.cell.com/cell-genomics/abstract/S2666-979X(22)00139-2

78. Abdelhalim A, Yilmaz O, Elshaikh Berair M, Torres T. A Narrative Review of the OX40-OX40L Pathway as a Potential Therapeutic Target in Atopic Dermatitis: Focus on Rocatinlimab and Amlitelimab. *Dermatol Ther (Heidelb)*. 2024;14(12):3197-3210. doi:10.1007/s13555-024-01308-8

79. Muehlschlegel S, Rordorf G, Bodock M, Sims JR. Dantrolene mediates vasorelaxation in cerebral vasoconstriction - A Case Series. *Neurocrit Care*. 2009;10(1):116-121. doi:10.1007/s12028-008-9132-5

80. Harding JJ, Piha-Paul SA, Shah RH, et al. Antitumour activity of neratinib in patients with HER2-mutant advanced biliary tract cancers. *Nature Communications*. 2023;14(1):630. doi:10.1038/s41467-023-36399-y

81. Zeitelhofer M, Adzemovic MZ, Moessinger C, et al. Blocking PDGF-CC signaling ameliorates multiple sclerosis-like neuroinflammation by inhibiting disruption of the blood–brain barrier. *Scientific Reports*. 2020;10(1):22383. doi:10.1038/s41598-020-79598-z

82. Adzemovic MZ, Zeitelhofer M, Eriksson U, Olsson T, Nilsson I. Imatinib Ameliorates Neuroinflammation in a Rat Model of Multiple Sclerosis by Enhancing Blood-Brain Barrier Integrity and by Modulating the Peripheral Immune Response. doi:10.1371/journal.pone.0056586

83. Tarimah K, Zulkarnaen DA, Widiastuti M, et al. Neuroprotective Effects of Magnesium Sulphate in Anesthesia and Neurocritical Care Settings: A Systematic Review. *F1000Research 2025 14:1318*. Preprint posted online November 26, 2025. doi:10.12688/f1000research.171465.1

84. Beck R, Verrax J, Dejeans N, Taper H, Calderon PB. Menadione Reduction by Pharmacological Doses of Ascorbate Induces an Oxidative Stress That Kills Breast Cancer Cells. *Int J Toxicol*. 2009;28(1):33-42. doi:10.1177/1091581809333139

85. Hong Y, Peng J, Chen Q, et al. Comparison of neoadjuvant chemotherapy response and prognosis among pegylated liposomal doxorubicin, epirubicin and pirarubicin in HR ⩽ 10%/HER2-negative breast cancer: an exploratory real-world multicentre cohort study. *Ther Adv Med Oncol*. 2024;16:17588359241279695. doi:10.1177/17588359241279695

86. Thorn CF, Oshiro C, Marsh S, et al. Doxorubicin pathways: pharmacodynamics and adverse effects. *Pharmacogenet Genomics*. 2011;21(7):440-446. doi:10.1097/FPC.0b013e32833ffb56

87. Dominguez-Mozo MI, López-Mecández D, Villar LM, et al. Short-chain fatty acids in multiple sclerosis: Associated with disability, number of T2 lesions, and inflammatory profile. *Annals of Clinical and Translational Neurology*. 2025;12(3):478-490. doi:10.1002/acn3.52259

88. Watanabe K, Taskesen E, Bochoven A van, Posthuma D. Functional mapping and annotation of genetic associations with FUMA. *Nature Communications*. 2017;8(1). doi:10.1038/s41467-017-01261-5

89. Soler-Sáez I, Gómez-Cabañes B, Grillo-Risco R, et al. Single cell landscape of sex differences in the progression of multiple sclerosis. *bioRxiv (Cold Spring Harbor Laboratory)*. Published online June 2024. doi:10.1101/2024.06.15.599139

90. Konuma T, Ogawa K, Okada Y. Integration of genetically regulated gene expression and pharmacological library provides therapeutic drug candidates. *Hum Mol Genet*. 2021;30(3-4):294-304. doi:10.1093/hmg/ddab049

91. Douville P, Martel AR, Talbot J, Desmeules S, Langlois S, Agharazii M. Impact of age on glomerular filtration estimates. *Nephrol Dial Transplant*. 2009;24(1):97-103. doi:10.1093/ndt/gfn473

92. Manouchehrinia A, Piehl F, Hillert J, et al. Confounding effect of blood volume and body mass index on blood neurofilament light chain levels. *Ann Clin Transl Neurol*. 2020;7(1):139-143. doi:10.1002/acn3.50972
